# Genomic epidemiology and evolution of rhinovirus in western Washington State, 2021-22

**DOI:** 10.1101/2024.02.13.24302773

**Authors:** Stephanie Goya, Seffir T. Wendm, Hong Xie, Tien V. Nguyen, Sarina Barnes, Rohit R. Shankar, Jaydee Sereewit, Kurtis Cruz, Ailyn C. Pérez-Osorio, Margaret G. Mills, Alexander L Greninger

## Abstract

**Background:** Human rhinoviruses (RV) primarily cause the common cold, but infection outcomes vary from subclinical to severe cases, including asthma exacerbations and fatal pneumonia in immunocompromised individuals. To date, therapeutic strategies have been hindered by the high diversity of serotypes. Global surveillance efforts have traditionally focused on sequencing VP1 or VP2/VP4 genetic regions, leaving gaps in understanding RV true genomic diversity.

**Methods:** We sequenced 1,003 RV genomes from nasal swabs of symptomatic and asymptomatic individuals to explore viral evolution during two epidemiologically distinct periods in Washington State: when the COVID-19 pandemic affected the circulation of other seasonal respiratory viruses except for RV (February – July 2021), and when the seasonal viruses reemerged with the severe RSV and influenza outbreak (November-December 2022). We constructed maximum likelihood and BEAST-phylodynamic trees to characterize intra-genotype evolution.

**Results:** We detected 100 of 168 known genotypes, identified two new genotypes (A111 and C59), and observed inter-genotypic recombination and genotype cluster swapping from 2021 to 2022. We found a significant association between the presence of symptoms and viral load, but not with RV species or genotype. Phylodynamic trees, polyprotein selection pressure, and Shannon diversity revealed co-circulation of divergent clades within genotypes with high amino acid constraints throughout polyprotein.

**Discussion:** Our study underscores the dynamic nature of RV genomic epidemiology within a localized geographic region, as more than 20% of existing genotypes within each RV species co-circulated each month. Our findings also emphasize the importance of investigating correlations between rhinovirus genotypes and serotypes to understand long-term immunity and cross-protection.

## Background

Human rhinoviruses (RV) are a primary etiology of the common cold, causing 8.9 – 19 % of total acute upper respiratory infections (ARI) and 4 – 6.1% of asymptomatic respiratory infections [1–4]. A recent study of RV socioeconomical burden in New York City found that 23.5% of the individuals with ARIs associated with RV missed at least 1 day of work or school (1). RV can also cause lower respiratory tract infections in immunocompromised patients with high mortality rate [5,6], and are a major contributor of exacerbations of asthma [7–9], cystic fibrosis [10] and chronic obstructive pulmonary disease [11,12]. While 100 different RV serotypes were described by 1987 [13], and the first complete RV genome was published in 1984 [14], fewer than 500 complete RV genomes were available in public databases at time of initiation of this study (January 2021), meaning most RV genotypes had fewer than 5 genomes available. Despite its more widespread disease burden, RV genomic data pale in comparison to the 163,000 influenza virus genomes or 11,500 respiratory syncytial virus (RSV) genomes currently available in GISAID or INSDC databases.

RV are non-enveloped viruses with a single-stranded positive-sense genome of around 7,300 nucleotides in length. The RV genome is organized into a single polyprotein that undergoes autocatalytically-based and protease-based cleavage to produce 4 capsid proteins (VP1 – VP4) and 7 non-structural proteins [15]. Global RV molecular epidemiology is based on phylogenetic association of VP1 genetic sequence, which allowed to classify three RV species (RV-A, RV-B, and RV-C), subclassified into genotypes (81 RV-A, 32 RV-B, and 55 RV-C genotypes) [16]. The co-circulation of many highly diverse genotypes, considered antigenically distinct, hampers the development of an effective broadly reactive RV vaccine [17].

Here, using a set of more than one thousand newly generated RV genomes constituting more than half of publicly available RV genomes, we describe the epidemiology and evolution of RV infections from symptomatic and asymptomatic individuals in Puget Sound region (Washington State, USA) during two distinct epidemiological time periods of the COVID-19 pandemic [18–22].

## Results

### Demographic and clinical characteristics of RV infections

Our RV screening was based on a subset of SARS-CoV-2 negative nasal swab specimens collected at the UW COVID-19 community testing sites taken during February-July 2021 and November-December 2022. A total of 1,083 out of 10,050 nasal swab specimens (10.8%) were RV positive from February-July 2021, and 820 out of 10,656 samples (7.8%) from November-December 2022. The RV positive population consisted of 49% male persons and 57% between the ages of 18 to 65 years (Table 1).

**Table 1.**
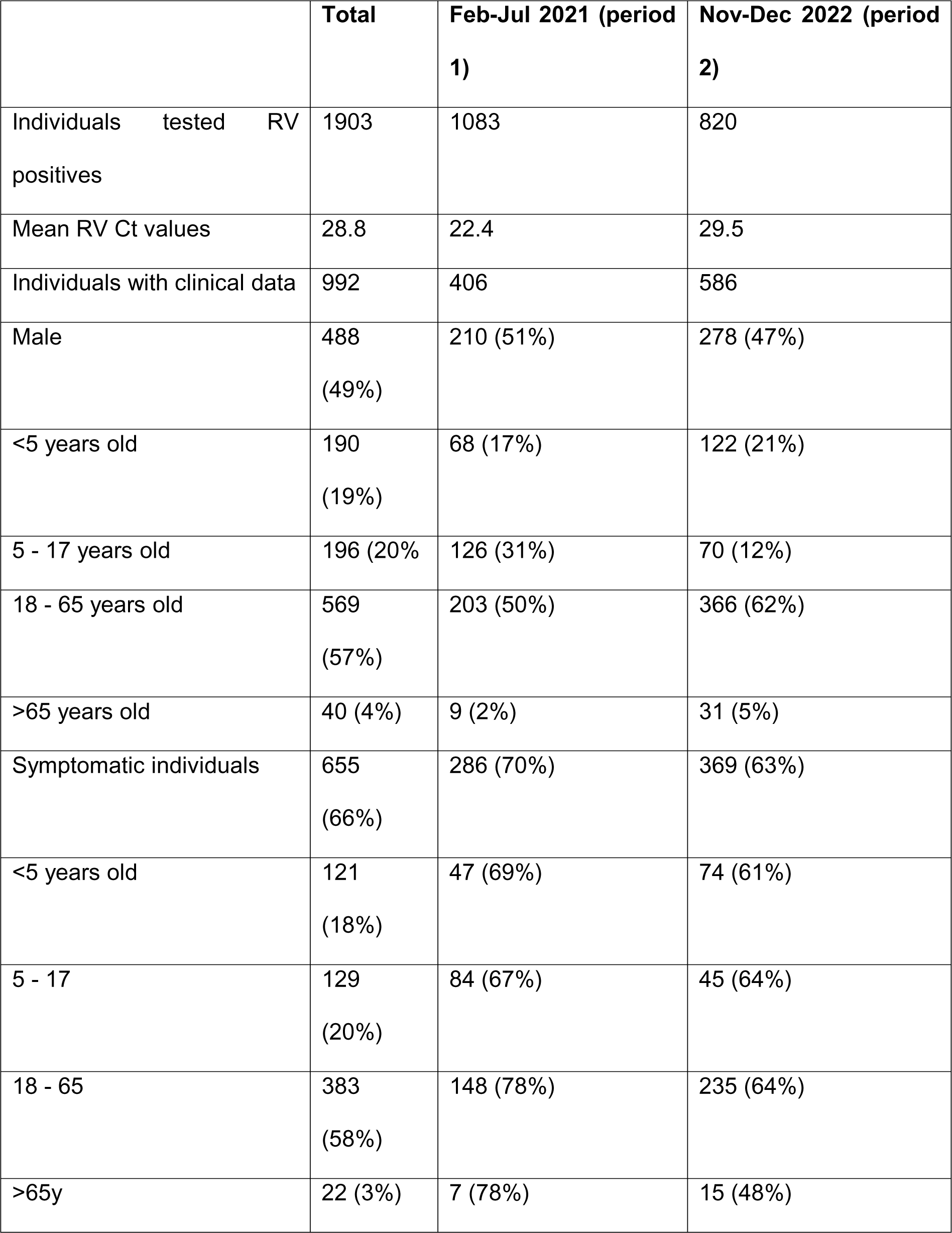
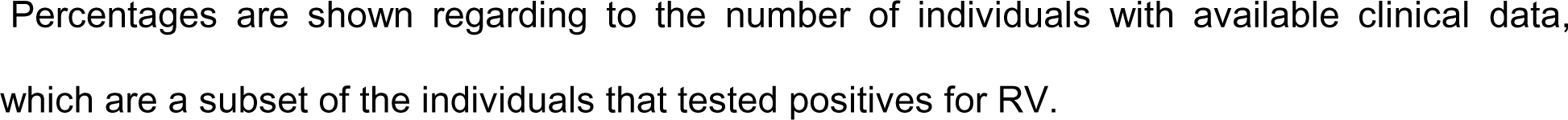
Demographic and virological characteristic of RV-positive individuals.

|  | **Total** | **Feb-Jul 2021 (period 1)** | **Nov-Dec 2022 (period 2)** |
| --- | --- | --- | --- |
| Individuals tested RV positives | 1903 | 1083 | 820 |
| Mean RV Ct values | 28.8 | 22.4 | 29.5 |
| Individuals with clinical data | 992 | 406 | 586 |
| Male | 488<br>(49%) | 210 (51%) | 278 (47%) |
| <5 years old | 190<br>(19%) | 68 (17%) | 122 (21%) |
| 5 - 17 years old | 196 (20%) | 126 (31%) | 70 (12%) |
| 18 - 65 years old | 569<br>(57%) | 203 (50%) | 366 (62%) |
| >65 years old | 40 (4%) | 9 (2%) | 31 (5%) |
| Symptomatic individuals | 655<br>(66%) | 286 (70%) | 369 (63%) |
| <5 years old | 121<br>(18%) | 47 (69%) | 74 (61%) |
| 5 - 17 | 129<br>(20%) | 84 (67%) | 45 (64%) |
| 18 - 65 | 383<br>(58%) | 148 (78%) | 235 (64%) |
| >65y | 22 (3%) | 7 (78%) | 15 (48%) |

Percentages are shown regarding to the number of individuals with available clinical data, which are a subset of the individuals that tested positives for RV.

Community testing site prevalence data correlated with that of Seattle hospital-based respiratory virus PCR surveillance data, which showed a RV monthly positivity rate of 13-25% from February-July 2021 and 5-10% in November-December 2022 (Figure 1). These two periods were epidemiologically distinct for respiratory virus circulation, as hospital-based surveillance data during February-July 2021 yielded a 4% SARS-CoV-2 positivity rate with no detection of other seasonal respiratory viruses. During November-December 2022, SARS-CoV-2 had a 10% average positivity rate, while seasonal respiratory viruses strongly re-emerged with a 34% positivity rate for RSV in November 2022, and 30% for influenza virus in both November and December.

**Figure 1.**
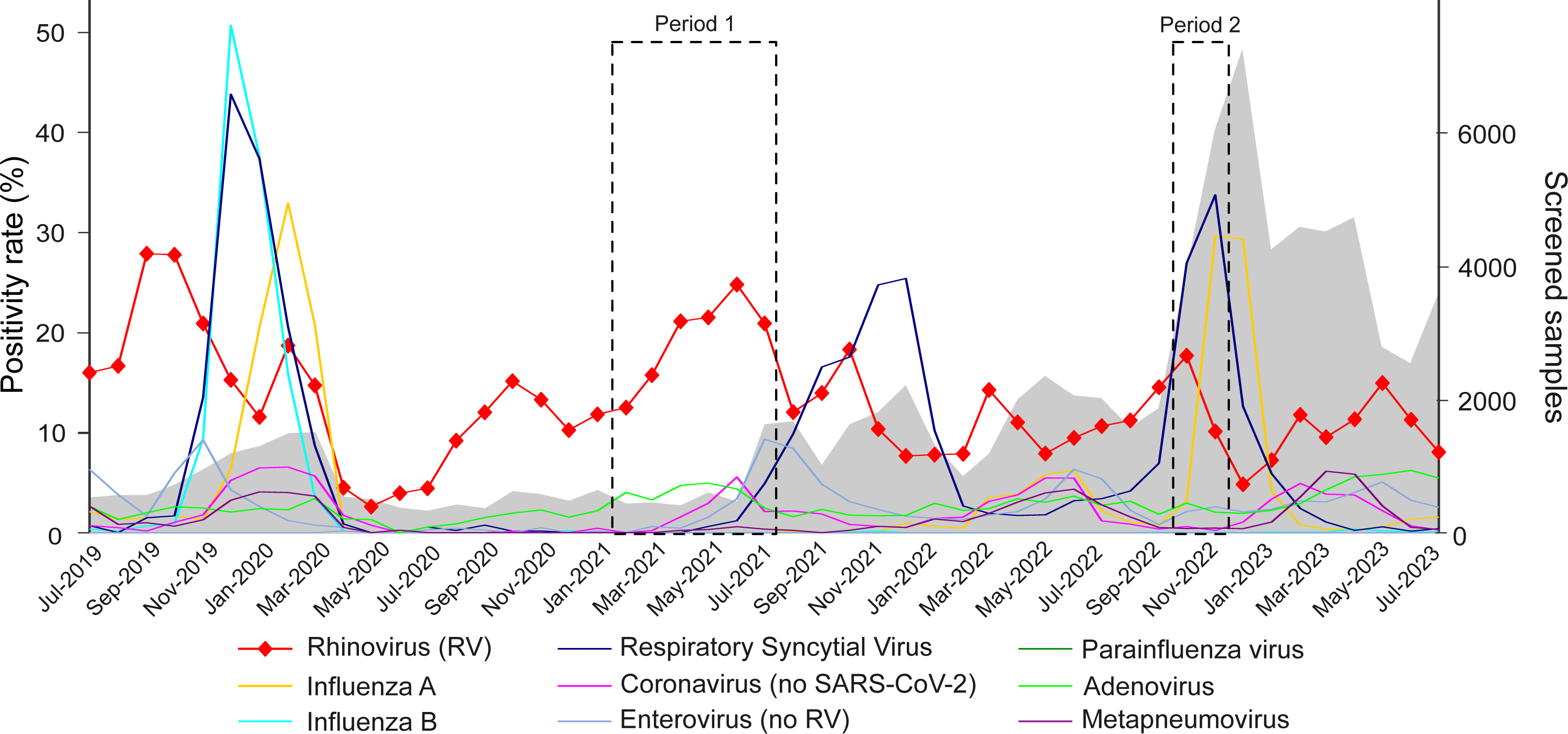
Seasonality of respiratory viruses in symptomatic patients in Washington State from 2019 to 2023. The positivity rate per month is detailed for different respiratory viruses. Rhinovirus is highlighted with red line including triangles. Grey shadow at the background of the plot informs the number of screened samples per month. The two studied periods of this work are denoted with dotted squares (P1= period 1, P2=period 2).

Among RV positives, persons aged 5 to 17 years exhibited significantly lower Ct values compared to individuals older than 18 years (Wilcoxon test, p-adj=1.0×10^-4^, Figure 2A). Symptomatic individuals comprised 66% of the RV positive population, including cough (35%), sore throat (23%), runny nose (17%), headache (13%), congestion (11%), and fever (9%) being the most common. Less commonly reported symptoms (<5% of cases) included sneezing, body ache, fatigue, difficulty breathing, and vomiting. Individuals reporting respiratory symptoms had lower Ct values than the asymptomatic individuals (Kruskal-Wallis test, p=2.1×10^-11^, Figure 2B). RV positives were slightly more likely to be symptomatic in 2021 compared to 2022 (70% vs. 63%) (Table 1). No association was found between the age of the individuals and the presence of symptoms (Figure 2C).

**Figure 2.**
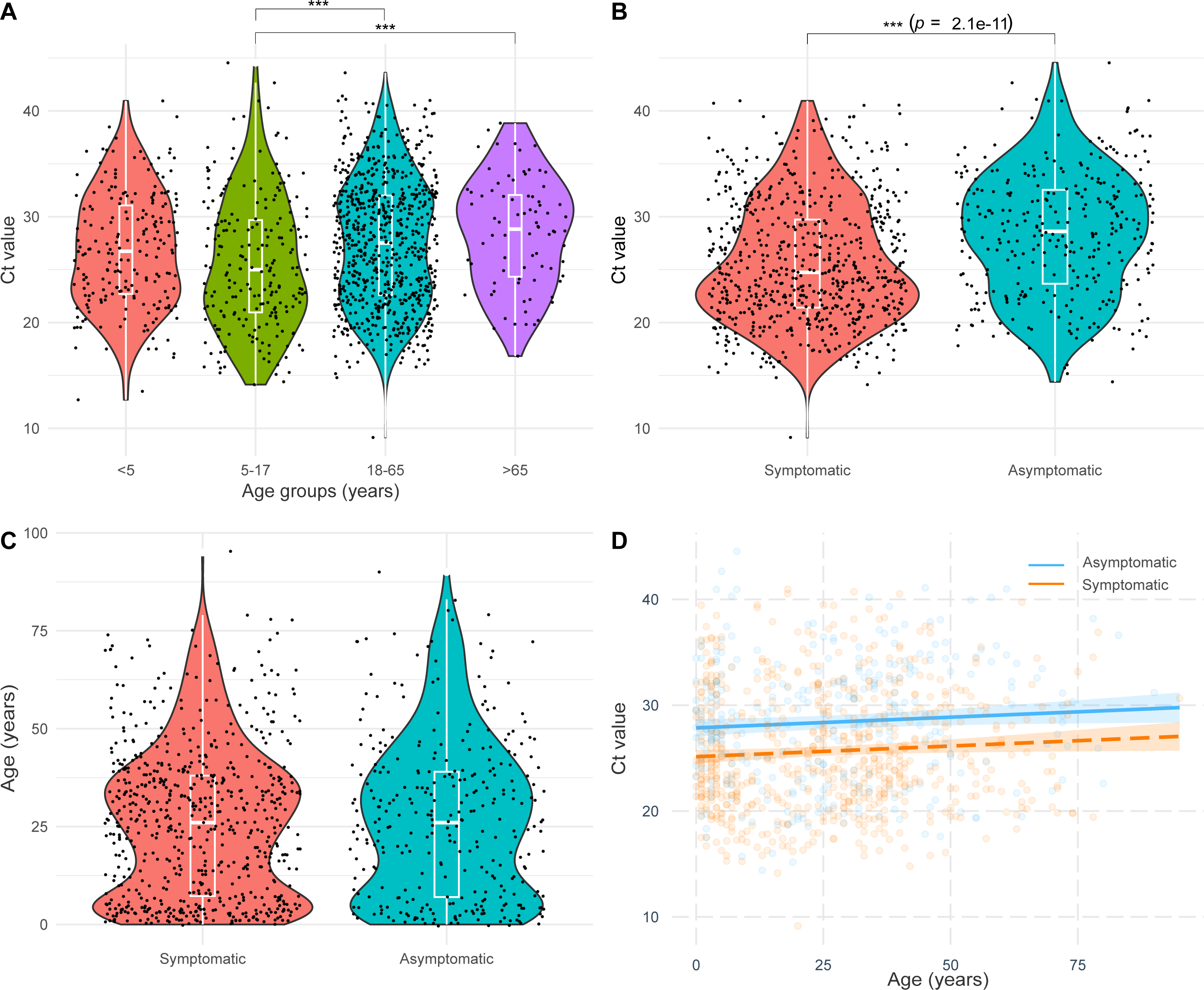
RV infections and relationship with the individuals’ age group and the symptoms presence. The association between the age of the individuals, presence of respiratory symptoms and the Ct value was evaluated. Violin plots illustrating the distribution of cases for Ct value and stratified age group (A), Ct value and symptoms presence (B) and non-stratified age and symptoms presence (C). Black dots represent individuals. Within the violin plot, a boxplot shows the median, interquartile range and 95% confidence interval. Kruskal-Wallis tests was used to assess the association between variables, but also Wilcoxon test for the age groups pairwise analysis. Supported (p value <0.001) differences are denoted with *** and includes details on the Kruskal-Wallis statistical test results. (D) Multivariate regression model including the Ct value, individuals’ age, and symptoms presence. Dots indicate individuals, colored by the symptom’s presence, same as the linear regression and the 95% confidence interval. Details on the multivariate prediction model are detail at the bottom of the plot.

Older individuals were associated with lower viral loads when adjusted for the presence of symptoms (ANOVA, p=0.03) (Figure 2D). Individuals with RV Ct value <25 had a significantly higher likelihood of reporting respiratory symptoms than those with Ct value >25 (Wald odds ratio= 1.63, 95% confidence limits: 1.03-2.59).

### Epidemiological and geographical characterization of RV species

We first characterized RV epidemiology at the species level. Our metagenomic sequencing effort resulted in 1,003 complete or almost complete RV genomes (883 from 2021 and 120 from 2022), consisting of 588 RV-A, 119 RV-B, and 296 RV-C. Seasonal analysis revealed an overall shift from RV-C to RV-A prevalence from February to July 2021. Combined with the higher prevalence of RV-C during November and December 2022, our data indicate a higher relative prevalence of RV-A in spring and summer with RV-C circulating at higher levels in autumn and winter (Figure 3A) [23]. RV-B exhibited 9.7% of the total cases in 2021 and 27.5% in 2022.

**Figure 3.**
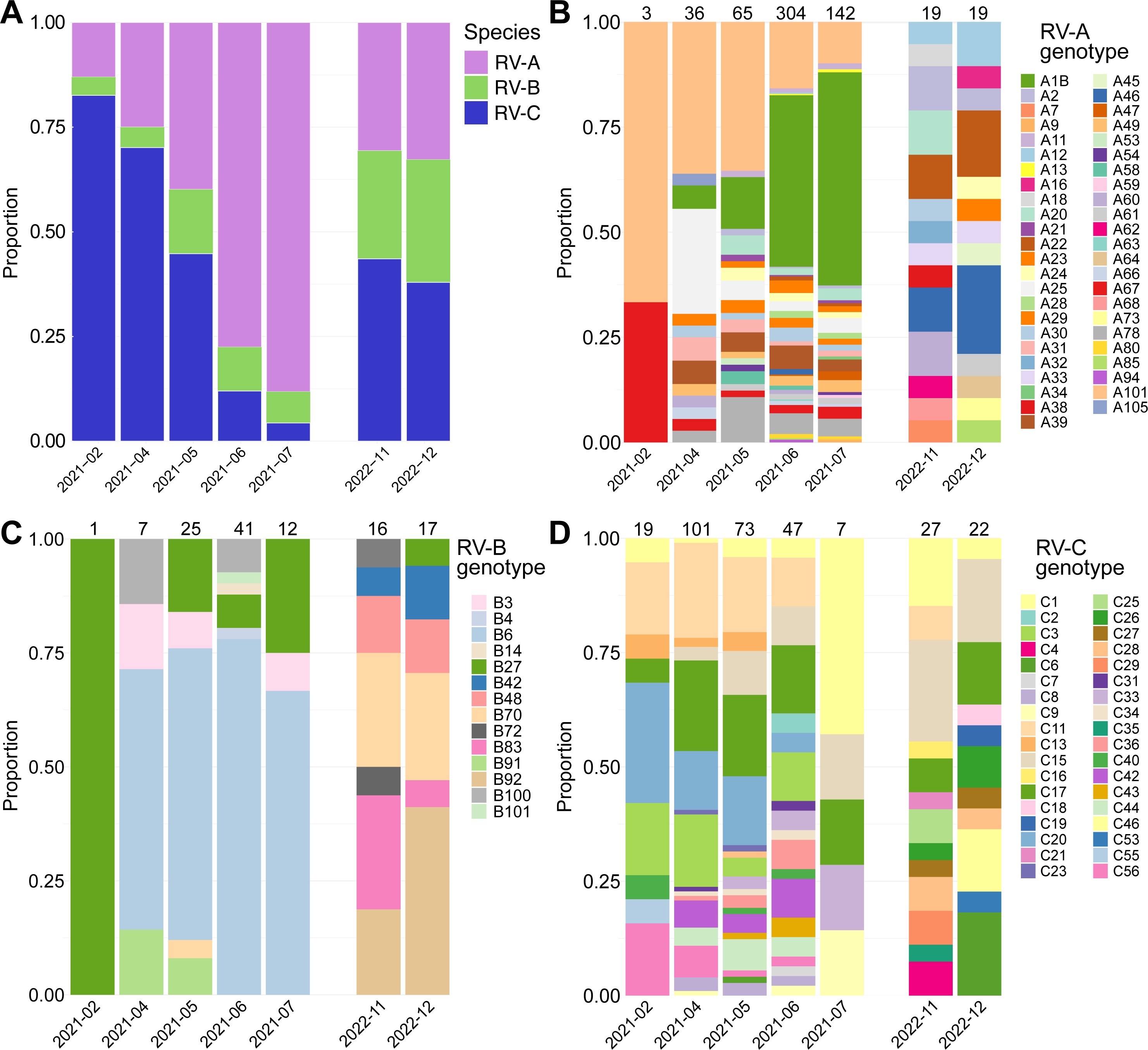
Abundance of RV species and genotypes within the sequenced genomes. The stacked bar-plot showed the proportion of RV species (A), genotypes within RV-A (B), genotypes within RV-B (C) and genotypes within RV-C (D) by studied month. For genotypes bar plots the total number of genomes per month is detailed above each column.

No difference in the presence of symptoms was observed by RV species (G-test of independence, p value>0.05). Children under 5 years old were more likely to be infected by RV-A or RV-C than RV-B (G-test of independence, p value = 2.02×10^-4^; CMLE odds ratio [95% confidence] RV-C vs RV-A=2.02 [1.19-3.47], RV-C vs RV-B=17.61 [2.86-723.92], RV-A vs RV-B=8.70 [1.39-361.16]). No tendency was found when analyzing RV species by non-stratified age.

We investigated if RV species distribution differed based on the location of COVID-19 community testing sites where specimens were collected. We found that the co-circulation, prevalence, and replacement of RV-A to RV-C was not geographically distinct during 2021, while no clear prevalence nor replacement was found in 2022, consistent with the overall seasonality (Figure 4).

**Figure 4.**
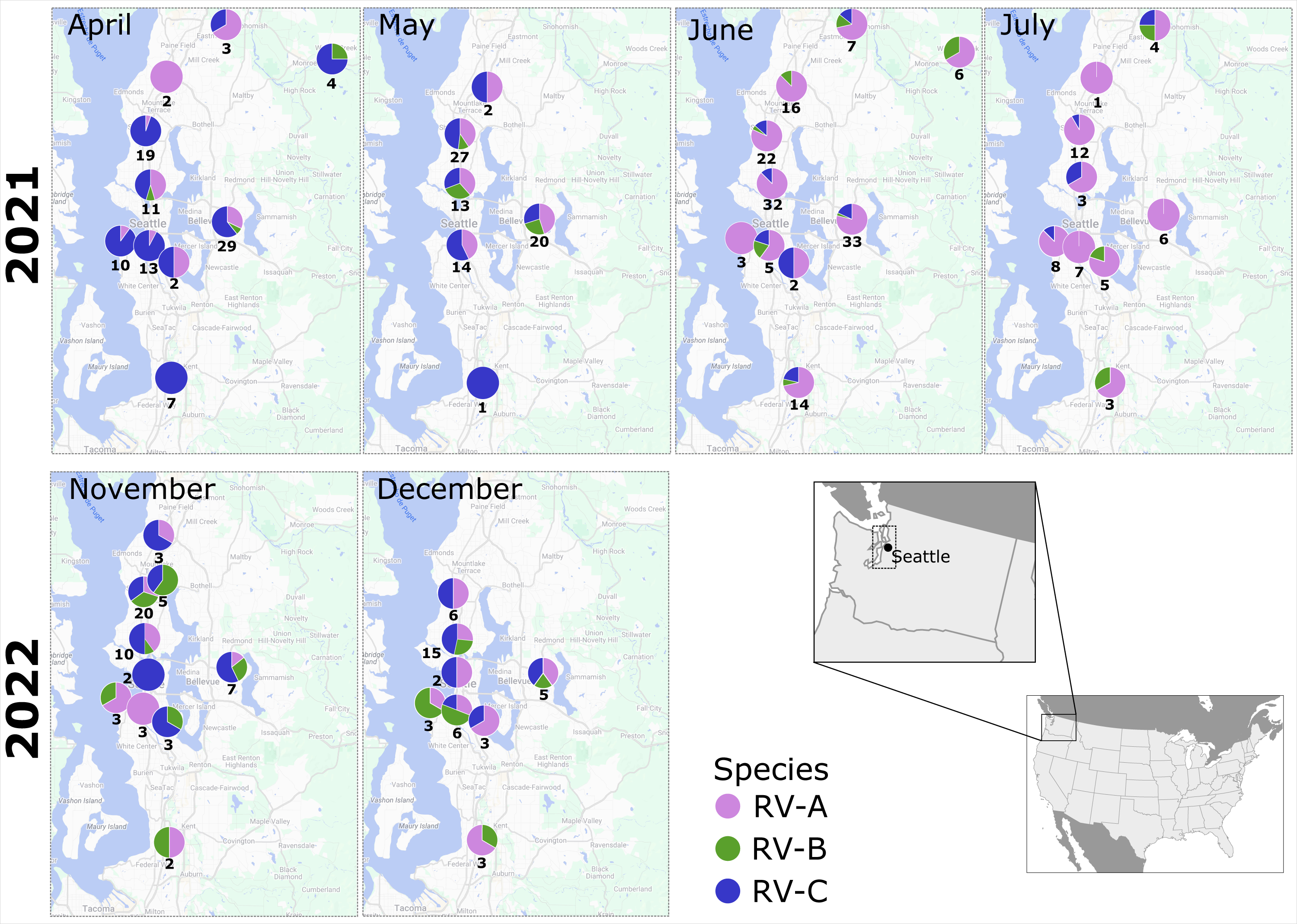
Geographic spread of RV species in Puget sound region. For each studied month the proportion of RV species (RV-A green, RV-B red and RV-C yellow) is mapped according to the University of Washington (UW) Medicine COVID-19 community testing sites where the individuals attended. Below each pie plot the total number of detected is informed. February 2021 is not shown due to the low number of cases available. At the bottom right a map highlighting the geographical location of Puget Sound is shown.

### RV genotype diversity in the Puget Sound region

We next characterized the RV diversity at the genotype level. Datasets of 1,327 RV-A, 314 RV-B, and 587 RV-C sequences were analyzed comprising all whole genomes available in NCBI Virus (October 2023), and VP1 reference sequences were included to represent RV-A and RV-C genotypes without available genomes. Within this dataset, 60% of sequences were collected in Washington State. To evaluate the continuity or reemergence of the genotype circulation in the Puget Sound region, 17 positive RV samples from the Seattle hospital-based respiratory virus surveillance, randomly sectioned from January to June 2023, were sequenced and included in the phylogenetic analyses. All complete genomes were trimmed to the VP1 region to proceed with the globally stablished genotyping classification [16].

We detected 47 RV-A genotypes, 14 RV-B genotypes, and 36 RV-C genotypes (Figure 3B-D, Figure 5A-B). A limited group of genotypes – A1B, A25, A39, A78, A101 in RV-A; B6 in RV-B; and C3, C11, C15, C17, C20 in RV-C – exhibited frequencies surpassing 1% of total cases though their prevalence varied throughout the studied time (Figure 5B). High RV-C prevalence from February to April 2021 was driven by four genotypes with limited change over time (C3, C11, C17, and C20), while RV-A dominance from May to July 2021 was accompanied by the replacement of A101 by A1B (Figure 3B-3D, Figure 5A). At the same time, B6 was the prevalent RV-B genotype during 2021. Remarkably, fifteen months later, most of the genotypes with high frequencies were not detected (Figure 5A).

**Figure 5.**
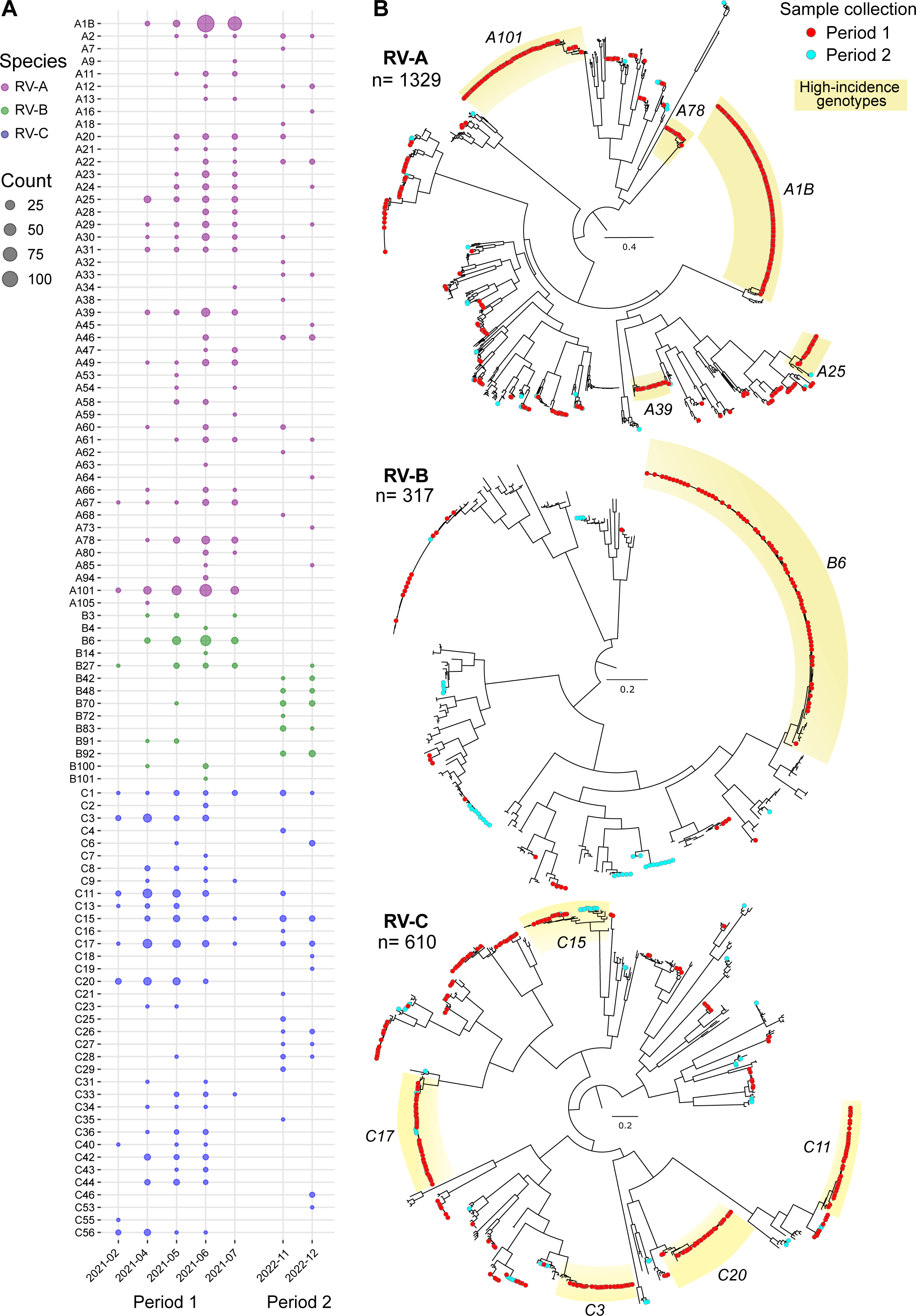
Genotyping of RV detected in Washington State in 2021 and 2022. A) Seasonality of RV genotypes detected within RV-A, RV-B and RV-C. The bubble chart details the number of cases (by the size of the circle) for each genotype per month of the studied period. Color in the circles denotes the RV species. B) Maximum likelihood tree with the VP1 nucleotide sequence of the cases studied in this work and reference sequences. Tree tips of sequences from this study are denoted with a red or light blue circle if were collected in period 1 or 2, respectively. The total number of sequences in the tree is informed below the rhinovirus specie. Phylogenetic clades of genotypes comprising more than 1% of the cases analyzed in this work are highlighted with yellow, including the name of the genotype in italics. Scale bar indicates substitution per site.

Rarefaction analysis indicated that the genotype richness was well characterized during 2021, while in 2022 genotypic coverage lagged despite our sampling efforts (Appendix, Supplementary Figure 1). Nonetheless, we covered more than 75% of estimated genotypic richness (Appendix). Based on the observed genotypic richness, Chao1 index calculation indicated the presence of up to 24 potentially unsampled genotypes in RV-A, 4 in RV-B and 17 in RV-C (Appendix).

### Novel RV-A and RV-C genotypes detected

We next examined the overall RV phylogenies for monophyletic clusters suggestive of novel genotypes (Supplementary Figure 2). Two monophyletic clusters exhibited distinctive patristic distances and further analysis indicated they had an average inter-genotype p-distance above the cutoff value of 0.13 (Supplementary Figure 2) [16]. We proposed them as genotypes A111 and C59. Genotype A111 comprised exclusively sequences from Washington State from 2017 to 2023, and C59 included sequences from Wisconsin from 2014 and Washington State from 2018 and 2023.

### Association of clusters of genotypes with clinical and demographic characteristics

The low number of cases for many of the detected RV genotypes impedes statistical association studies of clinical and epidemiological characteristics. Therefore, we evaluated whether phylogenetically-related groups of genotypes might demonstrate any such association. We analyzed principal component analysis (PCA) based on a polyprotein-based pairwise genetic distance matrix applying unsupervised hierarchical clustering. Genotypes were grouped into 4 clusters in RV-A, 5 clusters in RV-B, and 7 clusters in RV-C (Appendix, Supplementary Figure 3). While clusters showed no association with symptom, sex, or geographic location, they were associated with the age of the individuals. Specifically, in HRV-B, cluster 2 (genotypes B3, B72, B83, B92 and B100) was more related with older individuals (median 42 years old, interquartile range: 33-56 years old) cluster 1 (genotype B6, median 20 years old, interquartile range=10-32 years old) (Kruskal-Wallis test, p=0.04). In RV-A, cluster 4 (genotypes A7, A12, A20, A28, A45, A46, A58, A68, A78) was more associated with the group of individuals <5 and >65 years old, while cluster 2 (genotypes A2, A9, A11, A16, A18, A22-25, A29-34, A39, A47, A49, A54, A60-64, A66, A67, A73, A85, A94, A105) and cluster 3 (genotype A1B) were associated with the group individuals aged 5 to 64. While not statistically supported, RV-C cluster 4 (genotype C15) was more related with older individuals (mean 42 years old) compared to the other clusters (range of means: 21 – 29 years).

We also evaluated the seasonality of the cluster of genotypes and found that RV-A and RV-B clusters were clearly distinguished by the year of sample collection (p<0.05). Specifically, cluster 1 (A101) and 3 (A1B) in RV-A, and cluster 1 (B6) and 4 (B27) in RV-B were associated with detections in 2021 compared to other clusters. In the case of RV-C, the comparison by clusters did not result in a single supported association, however cluster 3 (C4, C9, C13, C19, C23, C25-27, C29, C33, C35, C36, C40, C46, C53, C55) and 4 (C15) were mainly detected in 2022, while cluster 7 (C20, C34) was mainly detected in 2021.

### Recombination between A105 and A21 genotypes within 3C protease gene

We next analyzed the complete genomes of the RV circulating in Puget Sound to examine for non-capsid changes. Interestingly, topological comparison of VP1 and 3D/RdRp phylogenetic trees revealed inconsistencies in a monophyletic clade comprised of four sequences from Washington State dating from 2016 and 2021. The clade was associated with A105 VP1 capsid sequence and A21 3D polymerase sequence (Figure 6A). Further analysis identified nucleotide 5,250 (95% confident interval: 5,216 – 5,271, GenBank accession number MZ268661), corresponding to amino acids 37-55 of the 3C protease, as the likely recombination breakpoint and was confirmed by all algorithms tested with RDP software (p<10^-13^) (Figure 6B).

**Figure 6.**
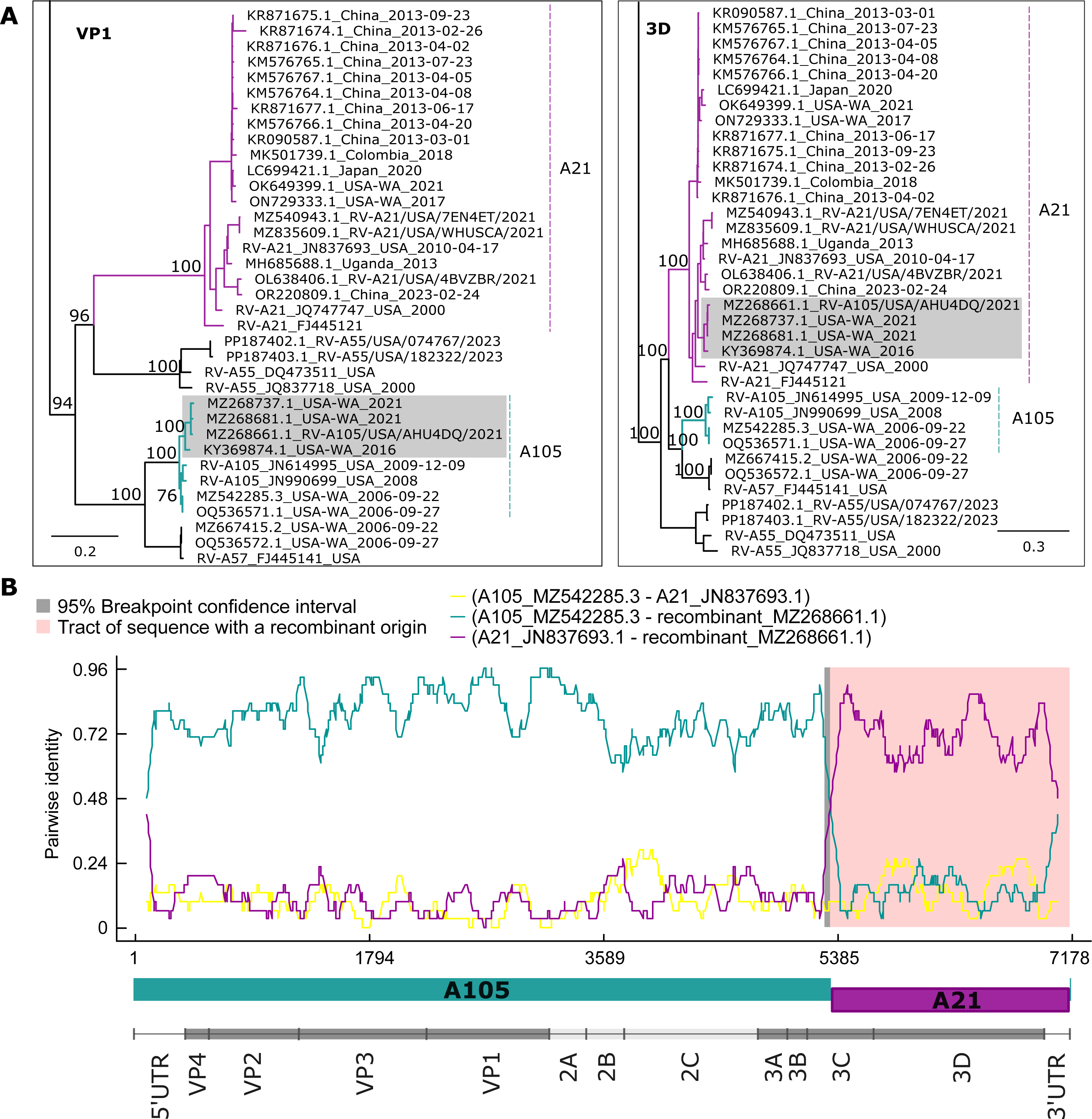
Recombination between RV-A105 and RV-A21 genotypes. A) Phylogenetic clades from the maximum likelihood trees with VP-1 and 3D nucleotide sequences. Branches are colored highlighting the inferred mayor (dark green) and minor (violet) parents for the recombinant clade (highlighted in grey). Bootstrap values of relevant nodes are indicated in the nodes. Scale bar indicates substitution per site. B) RDP pairwise identity plot between the sequences MZ542285 (A105, major parent), JN837693 (A21, minor parent) and MZ268661 (recombinant). The gray background denotes the 95% recombination breakpoint confidence interval and the pink background highlight the region with recombinant origin. At the bottom of the plot the representation of the parent combination and the RV genome organization following the same scale of nucleotide positions.

### Intra-genotypic RV evolution and variability

We reconstructed genome-based phylodynamic trees with the genomes available for the highly prevalent genotypes in this study (A1B, A25, A39, A78, A101 in RV-A; B6 in RV-B; and C3, C11, C15, C17, C20 in RV-C). Genotype A25 showed no temporal signal, and A39 failed to converge in BEAST analysis. The rest of genotypes showed consistently similar intra-genotype evolutionary (range: 2.15×10^-3^ - 4.54×10^-3^ substitutions/site/year, Figure 7), coinciding with inter-genotype evolutionary rates previously published [24,25]. The time of the most recent common ancestor (tMRCA) varied from 1982 to 2007. Ancestry of the RV circulating in Puget Sound in 2021 and 2022 showed that most of the sequences were closely associated into a few large clades, though some sequences were associated with clades that diverged decades ago. For example, sequences from the A101 genotype detected in 2021 were grouped into different clades sharing a MRCA from 1989, highlighting the evolutionary diversity within this genotype (Supplementary Figure 4). We wondered how the high evolutionary rate and divergence time might affect intra-genotypic variability at the amino acid level with possible antigenic consequences. Intra-genotype selection pressure estimation across the RV polyprotein unveiled overall strong negative selection spanning the entire open reading frame. However, positively selected sites for three genotypes were seen within the VP1 capsid protein, at positions 274 (genotype A1B), 43 (genotype A39) and 86 (genotype C15) (Appendix). Positively selected sites were also found within 3D (253 in genotype B6, 66 in genotype C3 and 133 in genotype C11), 3B (position 5, genotype C17) and 2A (position 58, genotype C3). In addition, the Shannon entropy calculation in the polyprotein revealed a single position (residue 86) in VP1 for the C15 genotype with moderate entropy (Supplementary Figure 5). Notably, all residues in all the other genotypes exhibited entropy values consistently below 1, indicating that all genotypes have a highly conserved polyprotein sequence.

## Discussion

In this study we analyzed the largest collection of newly sequenced rhinovirus (RV) genomes to date, comprising more than half of publicly available RV genomes in NCBI GenBank as of December 2023. We observed a high RV incidence during the studied period, even during November and December 2022 when the “tripledemic” of SARS-CoV-2, RSV, and influenza virus caused significant public health concern [26].

Our study also supports previous findings indicating that symptomatology is associated with higher viral loads than asymptomatic individuals. [27–29]. We observed a high incidence of RV-A in spring and summer, with RV-C predominating during the winter, consistent with prior reports [30]. However, unlike previous reports, we did not find age groups or symptomatology to be associated with specific RV species [3]. Our findings were limited to mild outpatient cases taken from community testing sites, and broader surveillance in severe infections could yield different results. Our study showed that the containment measures against the COVID-19 pandemic had no effect on the geographically spread of RV species, co-circulation, and replacement in the Puget Sound region, consistent with the high positivity rate seen for RV in hospital-based surveillance data. Further investigations globally are needed to assess if the seasonality we observed reflects a local or global circulation pattern, helping to determine if RV spread extends beyond strict border containment measures.

We detected 100 of 173 described RV genotypes, including 2 newly described genotypes and 3 other genotypes described in published manuscripts but not yet confirmed by the ICTV or *Picornaviridae* Study Group (Supplementary Figure 6). Overall, our work more than doubled the number of genomes available in the NCBI database for 65 RV genotypes. We detected the cocirculation of 30 RV-A genotypes in June 2021, that to the best of our knowledge is the highest number of genotypes observed in a single month for one RV species in a small geographic region. However, our sequencing efforts covered an estimated >70% of the genotypic richness, indicating there still were unseen genotypes. Although we have studied a distinctive epidemiological period that may have facilitated an increase in RV viral diversity in 2021, these results demonstrate the complexity of designing optimal RV molecular surveillance.

During our study period, 11 genotypes comprised >60% of studied RV cases. Interestingly, genotypes that predominated in 2021 were replaced in 2022. However, we did not perform serological analysis to determine whether population-level immunity to prevalent genotypes in 2021 defined the genotypic incidence the following year, or if it is related to another seasonality pattern. Prior work has shown that RV serotype-specific IgA in serum can last for at least one year, indicating that immunity is likely a contributing factor for shifting genotypic incidence [31].

Our genomic analysis detected a recombinant monophyletic clade between A105 and A21 genotypes with a breakpoint in the 3C protein region. A similar 3C recombination breakpoint has been reported between A76 and A56 genotypes [24,25]. This region of the genome is a hot spot for recombination events in poliovirus [32]. However, our A105-A21 recombination breakpoint was located upstream of the corresponding RNAse L inhibitory RNA secondary structure element, which was shown to be associated with higher rates of recombination in poliovirus [33,34]. Further investigation of the RNA secondary structure of RV may help to understand our findings.

We observed that RV co-circulating in the Puget Sound area from the same genotype diverged over a decade ago, though the polyprotein sequence exhibited a significant constraint on amino acid variability. Only three genotypes exhibited positive selection in single different sites of the VP1 protein, the major architectural component of the viral capsid and host-cell receptor attachment, while four other genotypes showed positively-selected sited in non-structural proteins. Further studies are needed to evaluate the biological impact of amino acid changes in these sites, but overall results suggest that the intra-genotypic amino acid variability is low, a promising fact for the development of an RV vaccine. However, this study emphasizes the importance of investigating correlations between genotypes and serotypes to understand long-term immunity and cross-protection. Recent research by Bochkov et al revealed cross-protection among some genetically similar genotypes in RV-A and RV-C species [35]. This fact was originally mentioned in 1982 when Cooney et al demonstrated cross-protection between 50 RV types, categorized them into 16 antigenic groups [36]. In our study, hierarchical clustering based on polyprotein genetic distances helped to associate some genotypes into a small number of clusters. We found statistical association of some clusters with the year of detection which might indicate a possible relationship with the long-term immunity interference. While this clustering does not necessarily define antigenic and serotypic relatedness, it did cluster together RV-A12, RV-20, and RV-A78 which have previously shown cross-neutralizing protection [35].

Our study has several limitations which may limit generalizability of findings, including the convenience sampling of remnant SARS-CoV-2 negative specimens, restriction of sampling to outpatient community testing sites, limited clinical metadata associated with high-throughput drive-thru testing, and sampling only during 2021-22 period within Puget Sound area. Nonetheless, the high volume of outpatient respiratory testing performed during the SARS-CoV-2 pandemic and omnipresent high co-circulation of RV made this study possible. Genomic recovery also requires specimens with higher viral loads, which may bias genomic epidemiological analyses such as to individuals with a symptomatic infection.

Overall, our deep look at RV genomic epidemiology during the 2021-2022 period in Puget Sound significantly increased the availability of genomic data for RV. We highlighted the seemingly unrestricted ability of RV species to co-circulate, including up to 30 RV-A genotypes in one month, despite public health containment measures that significantly limited co-circulation of other respiratory viruses. In addition, we showed the importance of update the RV surveillance to genomic approach to monitor recombination, and the continued need to examine antigenic and serotypic relatedness of RV genotypes that may reveal relationships with viral seasonality and vaccine development.

## Materials and methods

### Clinical samples and data collection

SARS-CoV-2-negative nasal swabs collected from symptomatic and asymptomatic individuals at University of Washington (UW) Medicine COVID-19 community testing sites were screened for RV by RT-qPCR (Appendix). Individuals declaring at least one respiratory symptom at time of collection were defined as symptomatic. This study was approved the UW Medicine Institutional Review Board with a consent waiver (STUDY00000408).

### NGS and assembly of viral consensus genomes

RNA from nasal swabs with RV Ct value below 33 were sequenced by metagenomic next-generation sequencing as described previously [37]. Twelve samples with Ct values 33-37 were also sequenced. Briefly, viral RNA was extracted with Quick-RNA Viral Kit (Zymo Research), double stranded cDNA synthesis was performed with random hexamers with SuperScript IV Reverse Transcriptase and Sequenase Version 2.0 DNA Polymerase (ThermoFisher Scientific) and purified with AMPure XP Magnetic Beads (Beckman Coulter). Library preparation was performed with Illumina DNA Prep, (S) Tagmentation kit (Illumina) and sequenced as 2 × 100-bp or 2 × 150-bp runs on a NovaSeq sequencer (Illumina). Consensus genomes were called using custom pipelines of mapping against references of all existing RV genomes from the International Committee on Taxonomy of Viruses (ICTV) as of March 2022 (Supplementary Material).

### Sequence analysis

RV complete genomes longer than 6,000 nt from human hosts were downloaded from in NCBI Virus (May 25, 2023) using the taxonomic IDs 147711 (RV-A), 147712 (RV-B), 463676 (RV-C). Partial genomes were downloaded from the *Picornaviridae* Study Group database to represent genotypes without complete genome available (https://www.picornastudygroup.com/). MEGA11 software was used to calculate the pairwise genetic p-distance for RV genotypes with 1,000 replicates bootstrapping [38]. Alignment-based recombination analysis was performed with RDP4 software using the RDP, GENECONV, Bootscan, Maximum Chi Square, Chimaera, SiScan, and 3SEQ methods with default settings [39]. Comparative co-phylogeny of VP1 and 3D regions was performed using ‘cophylo’ tool from the ‘phytools’ package in R [40].

### Statistical analyses

Rarefaction curves and the coverage-based extrapolation curves were calculated to estimate the genotypic diversity using the packages ‘vegan’ and ‘iNEXT’ in RStudio 2022.07.2 [41,42]. Association among categorical and continuous variables were evaluated with ANOVA, Kruskal-Wallis test, Wilcoxon test, G-test, or Wald odds ratio using AMR or epitools libraries in RStudio [43]. Principal component analysis (PCA) and hierarchical clustering (HC) based on pairwise genetic distances (p-distance) were performed using the packages ‘FactoMineR’, ‘factoextra’ and ‘eclust’ in RStudio (Supplementary Material). R markdown code is available at https://github.com/greninger-lab/HRV_epidemiology.

### Phylogenetic analyses

Sequences were aligned with MAFFT v7.490 and visualized with Aliview v1.28 [44,45]. Maximum likelihood trees were inferred with IQ-TREE v2.1 using ModelFinder to select the substitution model, and SH-aLRT test (1,000 replicates) and UFBoot2 method (1,000 replicates) to evaluate reliability of phylogenetic clades [46,47]. Comparative analysis of the genotyping procedure with maximum likelihood instead of neighbor joining inferences are detailed in Appendix.

Temporal signal was assessed with TempEst v1.5.3 [48]. The evolutionary rate was inferred with BEAST2 package v2.7.5 using an optimized relaxed clock and the tree priors selected according to the Nested Sampling test [49,50]. The convergence of the BEAST inference was assessed from the estimations of the Effective Sampling Size (ESS) and the highest posterior density interval (95% HPD) after a 10% burn-in using Tracer v1.7. TreeAnnotator was used to summarize the information from the sampled trees onto a single tree (the maximum clade credibility tree, MCCT).

## Data Availability

Sequencing data is available in NCBI BioProject PRJNA1066815

https://github.com/greninger-lab/HRV_epidemiology

## Sequence data availability

FASTQ and RV consensus genome data are available in NCBI BioProject PRJNA1066815 (SRA and GenBank accession numbers in Supplementary Table 1).

## Acknowledgments.

We thank the excellent technical support of Nathan Breit and Carlos Avendaño. This research received no specific funding and was supported by departmental funds.

## References

1. Hansen C, Perofsky AC, Burstein R, et al. Trends in Risk Factors and Symptoms Associated With SARS-CoV-2 and Rhinovirus Test Positivity in King County, Washington, June 2020 to July 2022. JAMA Netw Open. 2022; 5(12):e2245861.

2. Chow EJ, Casto AM, Roychoudhury P, et al. The Clinical and Genomic Epidemiology of Rhinovirus in Homeless Shelters-King County, Washington. J Infect Dis. 2022; 226(Suppl 3):S304–S314.

3. Zlateva KT, Rijn AL van, Simmonds P, et al. Molecular epidemiology and clinical impact of rhinovirus infections in adults during three epidemic seasons in 11 European countries (2007-2010). Thorax. 2020; 75(10):882–890.

4. Giardina FAM, Piralla A, Ferrari G, Zavaglio F, Cassaniti I, Baldanti F. Molecular Epidemiology of Rhinovirus/Enterovirus and Their Role on Cause Severe and Prolonged Infection in Hospitalized Patients. Microorganisms. 2022; 10(4):755.

5. Makhsous N, Goya S, Avendaño CC, et al. Within-host rhinovirus evolution in upper and lower respiratory tract highlights capsid variability and mutation-independent compartmentalization. J Infect Dis. 2023; :jiad284.

6. Boonyaratanakornkit J, Vivek M, Xie H, et al. Predictive Value of Respiratory Viral Detection in the Upper Respiratory Tract for Infection of the Lower Respiratory Tract With Hematopoietic Stem Cell Transplantation. The Journal of Infectious Diseases. 2020; 221(3):379–388.

7. Grissell TV, Powell H, Shafren DR, et al. Interleukin-10 gene expression in acute virus-induced asthma. Am J Respir Crit Care Med. 2005; 172(4):433–439.

8. Atmar RL, Guy E, Guntupalli KK, et al. Respiratory tract viral infections in inner-city asthmatic adults. Arch Intern Med. 1998; 158(22):2453–2459.

9. Castillo JR, Peters SP, Busse WW. Asthma Exacerbations: Pathogenesis, Prevention, and Treatment. J Allergy Clin Immunol Pract. 2017; 5(4):918–927.

10. Wat D, Gelder C, Hibbitts S, et al. The role of respiratory viruses in cystic fibrosis. J Cyst Fibros. 2008; 7(4):320–328.

11. McManus TE, Marley A-M, Baxter N, et al. Respiratory viral infection in exacerbations of COPD. Respir Med. 2008; 102(11):1575–1580.

12. Sim YS, Lee JH, Lee EG, et al. COPD Exacerbation-Related Pathogens and Previous COPD Treatment. J Clin Med. 2022; 12(1):111.

13. Hamparian VV, Colonno RJ, Cooney MK, et al. A collaborative report: rhinoviruses--extension of the numbering system from 89 to 100. Virology. 1987; 159(1):191–192.

14. Stanway G, Hughes PJ, Mountford RC, Minor PD, Almond JW. The complete nucleotide sequence of a common cold virus: human rhinovirus 14. Nucleic Acids Res. 1984; 12(20):7859–7875.

15. Esneau C, Bartlett N, Bochkov YA. Chapter 1 - Rhinovirus structure, replication, and classification. In: Bartlett N, Wark P, Knight D, editors. Rhinovirus Infections. Academic Press; 2019. p. 1–23.

16. McIntyre CL, Knowles NJ, Simmonds P. Proposals for the classification of human rhinovirus species A, B and C into genotypically assigned types. J Gen Virol. 2013; 94(Pt 8):1791–1806.

17. Esneau C, Duff AC, Bartlett NW. Understanding Rhinovirus Circulation and Impact on Illness. Viruses. 2022; 14(1):141.

18. Goya S, Sereewit J, Pfalmer D, et al. Genomic Characterization of Respiratory Syncytial Virus during 2022-23 Outbreak, Washington, USA. Emerg Infect Dis. 2023; 29(4):865–868.

19. Varela FH, Sartor ITS, Polese-Bonatto M, et al. Rhinovirus as the main co-circulating virus during the COVID-19 pandemic in children. J Pediatr (Rio J). 2022; 98(6):579–586.

20. Champredon D, Bancej C, Lee L, Buckrell S. Implications of the unexpected persistence of human rhinovirus/enterovirus during the COVID-19 pandemic in Canada. Influenza Other Respir Viruses. 2022; 16(2):190–192.

21. Georgieva I, Stoyanova A, Angelova S, Korsun N, Stoitsova S, Nikolaeva-Glomb L. Rhinovirus Genotypes Circulating in Bulgaria, 2018-2021. Viruses. 2023; 15(7):1608.

22. Pendrey CG, Strachan J, Peck H, et al. The re-emergence of influenza following the COVID-19 pandemic in Victoria, Australia, 2021 to 2022. Eurosurveillance. European Centre for Disease Prevention and Control; 2023; 28(37):2300118.

23. Morobe JM, Nyiro JU, Brand S, et al. Human rhinovirus spatial-temporal epidemiology in rural coastal Kenya, 2015-2016, observed through outpatient surveillance. Wellcome Open Res. 2019; 3:128.

24. McIntyre CL, Savolainen-Kopra C, Hovi T, Simmonds P. Recombination in the evolution of human rhinovirus genomes. Arch Virol. 2013; 158(7):1497–1515.

25. Zhao P, Shao N, Dong J, et al. Genetic diversity and characterization of rhinoviruses from Chinese clinical samples with a global perspective. Microbiology Spectrum. American Society for Microbiology; 2023; 0(0):e00840–23.

26. Nirappil F. Covid, flu, RSV declining in hospitals as ‘tripledemic’ threat fades. Washington Post [Internet]. 2023 [cited 2024 Jan 8];. Available from: https://www.washingtonpost.com/health/2023/01/22/covid-declining-flu-rsv-tripledemic/

27. Sanchez-Codez MI, Moyer K, Benavente-Fernández I, Leber AL, Ramilo O, Mejias A. Viral Loads and Disease Severity in Children with Rhinovirus-Associated Illnesses. Viruses. Multidisciplinary Digital Publishing Institute; 2021; 13(2):295.

28. Gerna G, Piralla A, Rovida F, et al. Correlation of rhinovirus load in the respiratory tract and clinical symptoms in hospitalized immunocompetent and immunocompromised patients. J Med Virol. 2009; 81(8):1498–1507.

29. Granados A, Peci A, McGeer A, Gubbay JB. Influenza and rhinovirus viral load and disease severity in upper respiratory tract infections. Journal of Clinical Virology. 2017; 86:14–19.

30. Wildenbeest JG, Schee MP van der, Hashimoto S, et al. Prevalence of rhinoviruses in young children of an unselected birth cohort from the Netherlands. Clin Microbiol Infect. 2016; 22(8):736.e9–736.e15.

31. Barclay WS, Nakib W al-, Higgins PG, Tyrrell DA. The time course of the humoral immune response to rhinovirus infection. Epidemiol Infect. 1989; 103(3):659–669.

32. Runckel C, Westesson O, Andino R, DeRisi JL. Identification and Manipulation of the Molecular Determinants Influencing Poliovirus Recombination. PLOS Pathogens. Public Library of Science; 2013; 9(2):e1003164.

33. Dutkiewicz M, Stachowiak A, Swiatkowska A, Ciesiołka J. Structure and function of RNA elements present in enteroviral genomes. Acta Biochimica Polonica. 2016; 63(4):623–630.

34. Burrill CP, Westesson O, Schulte MB, Strings VR, Segal M, Andino R. Global RNA Structure Analysis of Poliovirus Identifies a Conserved RNA Structure Involved in Viral Replication and Infectivity. J Virol. 2013; 87(21):11670–11683.

35. Bochkov YA, Devries M, Tetreault K, et al. Rhinoviruses A and C elicit long-lasting antibody responses with limited cross-neutralization. J Med Virol. 2023; 95(8):e29058.

36. Cooney MK, Fox JP, Kenny GE. Antigenic groupings of 90 rhinovirus serotypes. Infect Immun. 1982; 37(2):642–647.

37. Greninger AL, Waghmare A, Adler A, et al. Rule-Out Outbreak: 24-Hour Metagenomic Next-Generation Sequencing for Characterizing Respiratory Virus Source for Infection Prevention. J Pediatric Infect Dis Soc. 2017; 6(2):168–172.

38. Stecher G, Tamura K, Kumar S. Molecular Evolutionary Genetics Analysis (MEGA) for macOS. Molecular Biology and Evolution. 2020; 37(4):1237–1239.

39. Martin DP, Murrell B, Golden M, Khoosal A, Muhire B. RDP4: Detection and analysis of recombination patterns in virus genomes. Virus Evol. 2015; 1(1):vev003.

40. Revell LJ. phytools: an R package for phylogenetic comparative biology (and other things). Methods in Ecology and Evolution. 2012; 3(2):217–223.

41. Oksanen J, Simpson GL, Blanchet FG, et al. vegan: Community Ecology Package version 2.6-4 from CRAN [Internet]. 2022 [cited 2023 Dec 26]. Available from: https://rdrr.io/cran/vegan/

42. Chao A, Gotelli NJ, Hsieh TC, et al. Rarefaction and extrapolation with Hill numbers: a framework for sampling and estimation in species diversity studies. Ecological Monographs. 2014; 84(1):45–67.

43. Sullivan KM, Dean A, Soe MM. OpenEpi: A Web-based Epidemiologic and Statistical Calculator for Public Health. Public Health Rep. 2009; 124(3):471–474.

44. Katoh K, Standley DM. MAFFT Multiple Sequence Alignment Software Version 7: Improvements in Performance and Usability. Mol Biol Evol. 2013; 30(4):772–780.

45. Larsson A. AliView: a fast and lightweight alignment viewer and editor for large datasets. Bioinformatics. 2014; 30(22):3276–3278.

46. Minh BQ, Schmidt HA, Chernomor O, et al. IQ-TREE 2: New Models and Efficient Methods for Phylogenetic Inference in the Genomic Era. Molecular Biology and Evolution. 2020; 37(5).

47. Hoang DT, Chernomor O, Haeseler A von, Minh BQ, Vinh LS. UFBoot2: Improving the Ultrafast Bootstrap Approximation. Molecular Biology and Evolution. 2018; 35(2):518–522.

48. Rambaut A, Lam TT, Max Carvalho L, Pybus OG. Exploring the temporal structure of heterochronous sequences using TempEst (formerly Path-O-Gen). Virus Evolution. 2016; 2(1):vew007.

49. Bouckaert R, Vaughan TG, Barido-Sottani J, et al. BEAST 2.5: An advanced software platform for Bayesian evolutionary analysis. PLOS Computational Biology. Public Library of Science; 2019; 15(4):e1006650.

50. Kalyaanamoorthy S, Minh BQ, Wong TKF, Haeseler A von, Jermiin LS. ModelFinder: fast model selection for accurate phylogenetic estimates. Nat Methods. Nature Publishing Group; 2017; 14(6):587–589.

