## Supplementary Material for "Genomic epidemiology and evolution of rhinovirus in western Washington State, 2021-22"

**Supplementary Methods**

*Clinical samples and data collection*

A symptomatic respiratory infection was considered when the individual declared at least one respiratory symptom at the time of sample collection. Individuals declaring no respiratory symptoms but a close contact with a COVID-19 positive individual or requesting confirmation before a trip were identified as asymptomatic.

*Rhinovirus RT-qPCR*

Viral RNA was extracted from nasal swabs using the MagNA Pure 96 System (Roche). Extracted RNA underwent to RT-qPCR for RV with AgPath-ID One-Step RT-PCR (Life Technologies) or Luna Universal Probe One-Step RT-qPCR Kit (New England Biolabs) according to manufacturer recommendation, after demonstrating high correlation of same-sample RV Ct values between two kits. Primers and probes targeting the 5´ UTR of the RV genome were previously published[1]:

Primer -Forward: 5´-CP[A] GCC [T]GC GTG GY-3´

Primer -Reverse: 5´-GAA ACA CGG ACA CCC AAA GTA-3´

Probe: 5´-TCC TCC GGC CCC TGA ATG YGG C-3´

P= pyrimidine derivative, [A]= LNA-dA (Locked Nucleic Acid, LNA), [T]= LNA-dT, and Y= C/T

After sequencing RV genomes, we examined sequences for any mutations in the region where the primers and probe annealed to check for changes in Ct values/viral load quantitation. We detected the following mismatches in the forward primer (the mismatch is highlighted in bold):

**G**PAGCCTGCGTGGY: RV-C19, RV-C55, RV-C35, RV-C20, RV-C26, RV-C33, RV-C34, RV-C36, RV-C46, RV-C53, RV-C54, RV-C9, RV-C59

CP**T**GCCTGCGTGGY: RV-C11, RV-C15, RV-C23, RV-C25, RV-C2, RV-C40

CPAGCC**C**GCGTGGY: RV-A101, RV-C33, RV-C36, RV-C46, RV-C53, RV-C9, RV-C21, RV-C28, RV-C18, RV-C42, RV-C56, RV-C7, RV-C8

**T**PAGCCTGCGTGGY: RV-B26, RV-B99, RV-B42, RV-B5, RV-B91, RV-B70, RV-B17, RV-B69, RV-B104, RV-B52, RV-B48, RV-B35, RV-C43

We detected the following mismatches in the reverse primer (the mismatch is highlighted in bold):

GAAACACGG**TT**ACCCAAAGTA: RV-C20, RV-C34

GAAACACGGACA**A**CCAAAGTA: RV-A59

We detected the following mismatches in probe (the mismatch is highlighted in bold):

TCCTCCGGC**T**CCTGAATGYGGC: RV-A59

TCCTCCGG**T**CCCTGAATGYGGC: RV-C20, RV-C34

To evaluate whether the mismatches regarding the sequences of primers and probe affected the sensitivity of the RT-qPCR assay we compared the Ct values of the genotypes with and without mismatches. Results are available at <https://github.com/greninger-lab/HRV_epidemiology>. The average Ct for each genotype was variable, but there were no pattern associating high Ct values with the genotypes carrying mutations in the primers or probe, indicating the Ct variability would be related with another factor.

*NGS and assembly of viral consensus genomes*

Viral consensus genomes were called using the custom pipelines <https://github.com/Paul-rk-cruz/HRV_Pipeline> and <https://github.com/greninger-lab/revica> to map NGS reads against references of all existing RV genomes from the International Committee on Taxonomy of Viruses (ICTV) as of March 2022.

*Phylogenetic analyses*

Sequences were aligned with MAFFT v7.490 and visualized with Aliview v1.28 to detect and correct alignment artifacts, mainly around nucleotide gaps regions [2,3]. Maximum likelihood trees were inferred with IQ-TREE v2.1 using SH-aLRT test (1,000 replicates) and UFBoot2 method (1,000 replicates) to evaluate reliability of phylogenetic clades [4,5]. Monophyletic clusters (clades) were considered as statistically supported when UFBoot2 value ≥ 90% and SH-aLRT ≥ 80%. Phylogenetic tree visualization was performed with Figtree v1.4.4 (<https://github.com/rambaut/figtree>). Temporal signal was assessed with TempEst v1.5.3 [6].

The evolutionary rate was inferred with BEAST2 package v2.7.5 using an optimized relaxed clock, the substitution model was selected with ModelFinder and the tree priors selected according to the Nested Sampling test [7,8]. The convergence of the BEAST inference was assessed from the estimations of the Effective Sampling Size (ESS) and the highest posterior density interval (95% HPD) after a 10% burn-in using Tracer v1.7. When required, LogCombiner was used to combine up to four runs, each of them with 50 million generations of Markov chain Monte Carlo (MCMC) with a sample frequency to obtain 10,000 sampled trees. TreeAnnotator was used to summarize the information from the sampled trees onto a single tree (the maximum clade credibility tree, MCCT). Tree clades from BEAST were considered statistically supported when the posterior probability was ≥0.8.

*Evaluation of RV genotype classification with neighbor joining and maximum likelihood inferences*

We used the VP1 reference sequences reported by ICTV and the Picornaviridae Study Group database (<https://www.picornastudygroup.com/>) to evaluate the reliability of genotyping despite the inference methodology used. Specifically, we compared the tree topology obtained with neighbor joining (NJ) and maximum likelihood.

NJ trees were constructed with MEGA11 (Molecular Evolutionary Genetics Analysis) software using p-distance method, uniform rates among sites and pairwise deletion of gaps/missing data [9]. We included 1,000 Bootstrap replicates to assess the reliability of phylogenetic clades.

Maximum likelihood trees were constructed with IQ-TREE v2.1, using ModelFinder to select the suitable nucleotide substitution model and SH-aLRT test (1,000 replicates) and UFBoot2 method (1,000 replicates) to evaluate reliability of phylogenetic clades [4,5,8]. Phylogenetic clades were considered as statistically supported when UFBoot2 value ≥ 90% and SH-aLRT ≥ 80%.

A total of 197 RV-A, 67 RV-B and 81 RV-C sequences were used for this analysis. The constructed trees are available at https://github.com/greninger-lab/HRV_epidemiology and the sequences names contain the NCBI GenBank accession number.

The comparison was evaluated with tanglegram plots (representation of the co-phylogeny in which the two phylogenetic trees are linked by the tips) constructed in RStudio with the package ‘phytools’ [10]. The trees were visually inspected with FigTree v1.4.4 (<https://github.com/rambaut/figtree>). Constructed trees and tanglegrams are available at https://github.com/greninger-lab/HRV_epidemiology.The clades association was consistent between different inferences for the three RV species, with minimal topological differences and a robust statistical support for genotype-defining nodes. Considering the results, we used IQ-TREE software (maximum likelihood inference) throughout our study since its suitability to deal with large datasets [4].

*Rarefaction and extrapolation curves to evaluate the RV genotype diversity (richness) covered in this study*

The coverage of RV genotypic diversity per year and month of sample collection was estimated with rarefaction curves with the packages ‘vegan’ and the coverage-based extrapolation curves with the package ‘iNEXT’ in RStudio [11,12]. Details on the code used is available in the R Markdown files RV_rarefaction.Rmd and RV_rarefaction.html at https://github.com/greninger-lab/HRV_epidemiology, including the data frames.

Rarefaction curves were calculated with a step size of 1. **Supplementary figure 1** shows the results of the RV genotypes per year of sample collection, and the curves of the genotypes per species and month of sample collection. In 2021 the genotype richness was well characterized denoted by a curve reaching the plateau, while during 2022 the curve still increased meaning that our sequencing depth was not enough, and more genotypes may be detected with more sequencing efforts (**Supplementary figure 1**). The evaluation per month and RV species detailed in more depth the situation, marked by the rarefaction curve proximate to the asymptote in some but not all RV species for a given month. For example, while in June 2021 the sampling size optimally describes the genotypic diversity of RV-A, in RV-B and RV-C species the genotypic richness is not totally represented.

In addition, estimation of genotype richness (Hill number q=0) was performed on the individual-based abundance data. Then the coverage-based extrapolation curve was performed. Bottom of **Supplementary figure 1** shows the genotype richness estimator per year and the estimation per RV species and month of sample collection, as a function of sample coverage. Solid lines of the curves represent rarefaction while dashed curves represent extrapolation beyond observed samples. The shaded areas highlight the 95% confidence intervals calculated from bootstrap. The results show that most of the times our sampling efforts covered more than 75% of estimated genotypic diversity (**Supplementary figure 1)**. We estimated the total number of genotypes per species for the months covering more than 75% diversity using the Chao1 index inference in ‘iNEXT’ package. The calculation indicated a range of 23-54 genotypes monthly co-circulating in RV-A, 3-11 genotypes in RV-B and 15-33 genotypes in RV-C (Supplementary table 1).

**Characterization of the new genotypes A111 and C59.**

During the phylogenetic analysis interpretation, three distinctive monophyletic clusters were observed with no close genotype reference sequence. The patristic distances (branch length) of the clades were equivalent to those differentiating genotypes, suggesting potential novel genotypes. Further analysis with recently new reported genotypes, but not confirmed by ICTV, identified an RV-B sequence from November 2022 as B107 genotype. Nevertheless, the other two clades did not have a close reference genotype. In **Supplemental Figure 2**, partial phylogenetic trees of RV-A (8A) and RV-C (8B) are shown with details about the supported clades in the nodes. Gray highlights the closest genotype/s including the genotype’s name at the left. Red boxes highlight the clade of the new genotypes, including the genotype’s name at the right. Scale bar indicates substitution per site.

At the right of each subtree, the evolutionary divergence, calculated as average VP1 p-distance between and within genotypes is informed in bold and the standard error estimation in blue and italics. Average VP1 p-distance between clades above 0.13 (<87% identity) indicates they are new genotypes.

**Principal component analysis (PCA) and hierarchical clustering**

We wondered if individual were associated with age group, gender, or respiratory symptoms of the individuals. For the genotypes with a number of detected cases allowing to apply a statistical analysis -A101, A1B, B6, and C11- no association was found (G-Test, p>0.05). Therefore, we evaluated whether a group of genotypes instead of independently were correlated using principal component analysis (PCA) based on the polyprotein-based pairwise genetic distance (p-distance) matrix as a categorization complexity between species and genotypes. The pairwise genetic distance (p-distance) matrix was calculated for each RV species with the package ‘ape’ in RStudio based on a nucleotide alignment of the polyprotein gene. PCA was calculated and evaluated with the package ‘FactoMineR’ and ‘factoextra’ in RStudio. Details about the pipeline used are available at https://github.com/greninger-lab/HRV_epidemiology. We analyze with hierarchical clustering (HC) whether unsupervised clustering (package ‘eclust’ in RStudio) of the data can be explained with the clinical and demographic characteristics of the individuals: age group, sex, symptoms presence and geographic location of the COVID-19 community testing sites. HC was calculated based on complete-linkage method and Euclidean distance, assessing the number of clusters with WSS (within sum of squares) and Silhouette score methods. HC grouped the sequences into 4 clusters in RV-A, 4 clusters in RV-B and 7 clusters in RV-C (**Supplemental Figure 3**, numbered clusters identified with colors and below each cluster the group of genotypes is detailed). The HC clustering showed no relation with the symptom presence, individual’s sex, or geographic location (detailed results available at https://github.com/greninger-lab/HRV_epidemiology). However, supported correlation with the age and RV-B HC clusters was found. Specifically, cluster 1 (genotypes B3, B72, B83, B92 and B100) have a higher incidence in individuals younger than 5 years old or older than 65 years old, while cluster 4 (genotype B6) was detected only in individuals from 5 to 64 years old (G-test of independence, p-value <0.05). Consistently, the analysis with non-stratified age showed supported differentiation between those clades (Kruskal-Wallis test, p value=0.04) (**Supplemental Figure 3**). The mean age for individuals within cluster 1 was 42 with a standard deviation of 19.1 and included the unique case of 4 years-old children included in the RV-B PCA analysis. Additionally, the mean age for cluster 4 was 20 with a standard deviation of 12.8. Similarly, a supported correlation between the HC clusters and the stratified age was detected in RV-A, but it was not supported during the analysis of non-stratified age (G-test of independence, p-value <0.05, **Supplemental Figure 3**). Precisely, HC cluster 2 (genotypes A2, A9, A11, A16, A18, A22-25, A29-34, A39, A47, A49, A54, A60-64, A66, A67, A73, A85, A94, A105) and cluster 4 (genotype A1B) were mainly related with individuals aged 5 to 64. However, HC cluster 3 (genotypes A7, A12, A20, A28, A45, A46, A58, A68, A78) were associated with individuals younger than 5 years old and older than 65 years old. RV-C showed no supported correlation for the studied variables, but a tendency in HC cluster 3 (genotype C15) with older individuals was observed (**Supplemental Figure 3**).

**Analysis of the intra-genotypic RV variability.**

Selection pressure within the RV polyprotein was calculated for each genotype independently. The datasets were nucleotide alignments of genomes from 2021 and 2022 collected in Washington State together with all the genomes from the same genotype available in public databases. The method used for the calculation was FUBAR to infer nonsynoymous (dN) and synonymous (dS) substitution rates on a per-site basis for the polyprotein coding alignment and the corresponding phylogeny within the online website <https://www.datamonkey.org/> [13]. Result of the selection pressure estimation per position and genotype are available at https://github.com/greninger-lab/HRV_epidemiology. Assuming that the selection pressure for each site is constant along the entire reconstructed phylogeny, FUBAR found strong negative selection spanning the polyprotein. However, positive selected sites for three genotypes were seen within the VP1 capsid protein, at positions 274 (genotype A1B), 43 (genotype A39) and 86 (genotype C15). Positive selected sites were also found within 3D (253 in genotype B6, 66 in genotype C3 and 133 in genotype C11), 3B (position 5, genotype C17) and P2-A (position 58, genotype C3).

Shannon entropy was calculated online at <https://www.hiv.lanl.gov/> for each prevalent genotype using the alignment of the open reading frame of the RV polyprotein translated with the standard genetic code. For this purpose, genomes from 2021 and 2022 collected in Washington State together with all the genomes from the same genotype available in public databases were analyzed. **Supplemental Figure 5** shows the Shannon entropy value as bars at each position of the entire polyprotein. Grey columns denote values below 1.3, meaning low variable positions in all the polyprotein alignment. The red column in position 86 in VP1 for genotype C15 denote a value above 1.3, therefore a moderately variable position.

**Bibliography**

1.	Weinberg GA, Schnabel KC, Erdman DD, et al. Field evaluation of TaqMan Array Card (TAC) for the simultaneous detection of multiple respiratory viruses in children with acute respiratory infection. J Clin Virol. **2013**; 57(3):254–260.

2.	Katoh K, Standley DM. MAFFT Multiple Sequence Alignment Software Version 7: Improvements in Performance and Usability. Mol Biol Evol. **2013**; 30(4):772–780.

3.	Larsson A. AliView: a fast and lightweight alignment viewer and editor for large datasets. Bioinformatics. **2014**; 30(22):3276–3278.

4.	Minh BQ, Schmidt HA, Chernomor O, et al. IQ-TREE 2: New Models and Efficient Methods for Phylogenetic Inference in the Genomic Era. Molecular Biology and Evolution. **2020**; 37(5).

5.	Hoang DT, Chernomor O, Haeseler A von, Minh BQ, Vinh LS. UFBoot2: Improving the Ultrafast Bootstrap Approximation. Molecular Biology and Evolution. **2018**; 35(2):518–522.

6.	Rambaut A, Lam TT, Max Carvalho L, Pybus OG. Exploring the temporal structure of heterochronous sequences using TempEst (formerly Path-O-Gen). Virus Evolution. **2016**; 2(1):vew007.

7.	Bouckaert R, Vaughan TG, Barido-Sottani J, et al. BEAST 2.5: An advanced software platform for Bayesian evolutionary analysis. PLOS Computational Biology. Public Library of Science; **2019**; 15(4):e1006650.

8.	Kalyaanamoorthy S, Minh BQ, Wong TKF, Haeseler A von, Jermiin LS. ModelFinder: fast model selection for accurate phylogenetic estimates. Nat Methods. Nature Publishing Group; **2017**; 14(6):587–589.

9.	Stecher G, Tamura K, Kumar S. Molecular Evolutionary Genetics Analysis (MEGA) for macOS. Molecular Biology and Evolution. **2020**; 37(4):1237–1239.

10.	Revell LJ. phytools: an R package for phylogenetic comparative biology (and other things). Methods in Ecology and Evolution. **2012**; 3(2):217–223.

11.	Oksanen J, Simpson GL, Blanchet FG, et al. vegan: Community Ecology Package version 2.6-4 from CRAN. 2022.

12.	Chao A, Gotelli NJ, Hsieh TC, et al. Rarefaction and extrapolation with Hill numbers: a framework for sampling and estimation in species diversity studies. Ecological Monographs. **2014**; 84(1):45–67.

13.	Murrell B, Moola S, Mabona A, et al. FUBAR: A Fast, Unconstrained Bayesian AppRoximation for Inferring Selection. Molecular Biology and Evolution. **2013**; 30(5):1196–1205.
