## Supplementary Figure 1 for "Genomic epidemiology and evolution of rhinovirus in western Washington State, 2021-22"

**Supplementary Figure 1. Rarefaction curves of the diversity of RV genotypes detected.**
The rarefaction curve plots represent the number of genomes sampled against the genotypic richness detected. The coverage-based rarefaction (solid lines) and extrapolation (dashed lines) curve plots are based on species richness (Hill's numbers q=0) and include the 95% confidence intervals as shaded areas based on 200 replication bootstrap method. For both analysis, rarefaction and coverage-based extrapolation, all RV species per year of sample collection and each RV species per month of sample collection were analyzed.

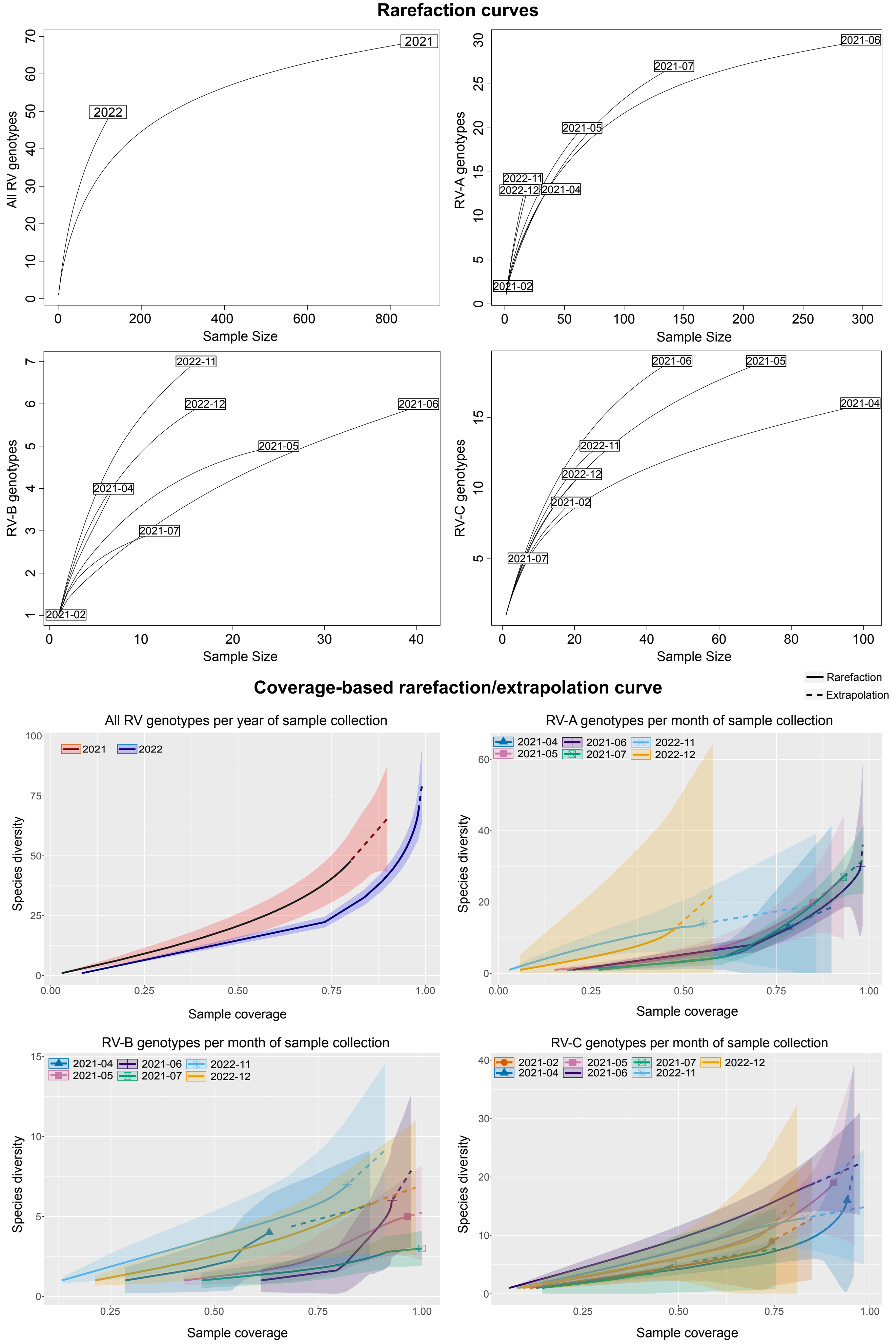
