## Supplementary Figure 2 for "Genomic epidemiology and evolution of rhinovirus in western Washington State, 2021-22"

**Supplementary Figure 2. Detection of the new genotypes A111 and C59.**

Characterization of the new genotypes A111 (A) and C59 (B). Partial phylogenetic trees show the monophyletic clade statistically supported by ultrafast bootstrap (with value above 90) indicated with a red square and the closest genotype/s highlighted in gray. Scale bar indicates substitution per site. At the right of each subtree, the evolutionary divergence (calculated as average p-distance) between and within genotype clades is informed in bold and the standard error estimation in blue italics.

**A**

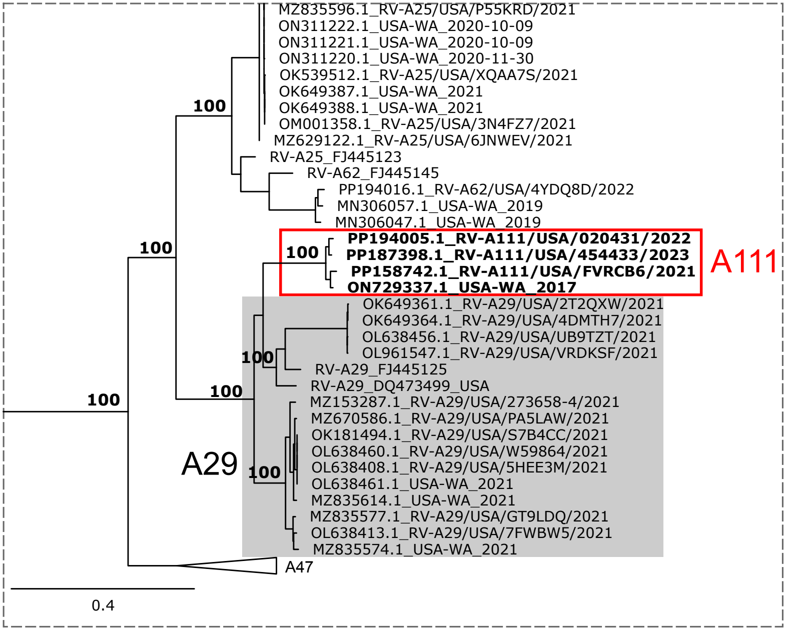


Average p-distance between clades

| | A111 | A29 |
|---|---|---|
| A111 | | *0.009* |
| A29 | **0.153** | |

Average p-distance within clade

| | | |
|---|---|---|
| A111 | **0.025** | *0.004* |
| A29 | **0.087** | *0.006* |

**B**

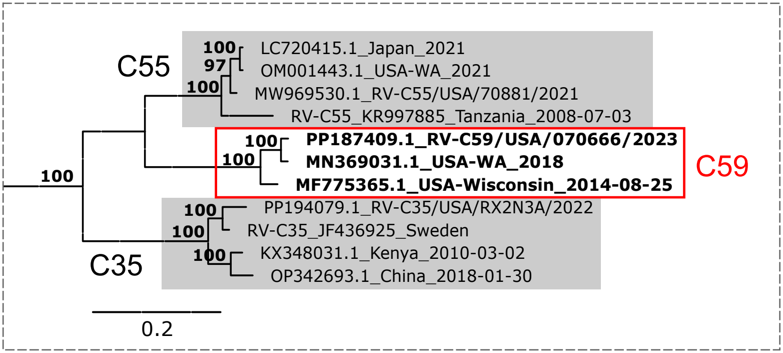


Average p-distance between clades

| | C59 | C35 | C55 |
|---|---|---|---|
| C59 | | *0.013* | *0.012* |
| C35 | **0.224** | | *0.013* |
| C55 | **0.197** | **0.223** | |

Average p-distance within clade

| | | |
|---|---|---|
| C59 | **0.052** | *0.007* |
| C35 | **0.078** | *0.007* |
| C55 | **0.057** | *0.007* |
