## Supplementary Figure 3 for "Genomic epidemiology and evolution of rhinovirus in western Washington State, 2021-22"

**Supplementary Figure 3. Hierarchical cluster analysis of the polyprotein-based pairwise genetic distance matrix.** Principal component analysis (PCA) and hierarchical clustering was calculated from the pairwise genetic distance based on the polyprotein sequence for each RV species. The number of clusters was defined by within-cluster sum of squares method. Each numbered cluster is highlighted with a colored squared including the detail of the genotypes clustering together. At the right of each dendrogram the analysis of association between age of the individuals and the clusters obtained is shown.

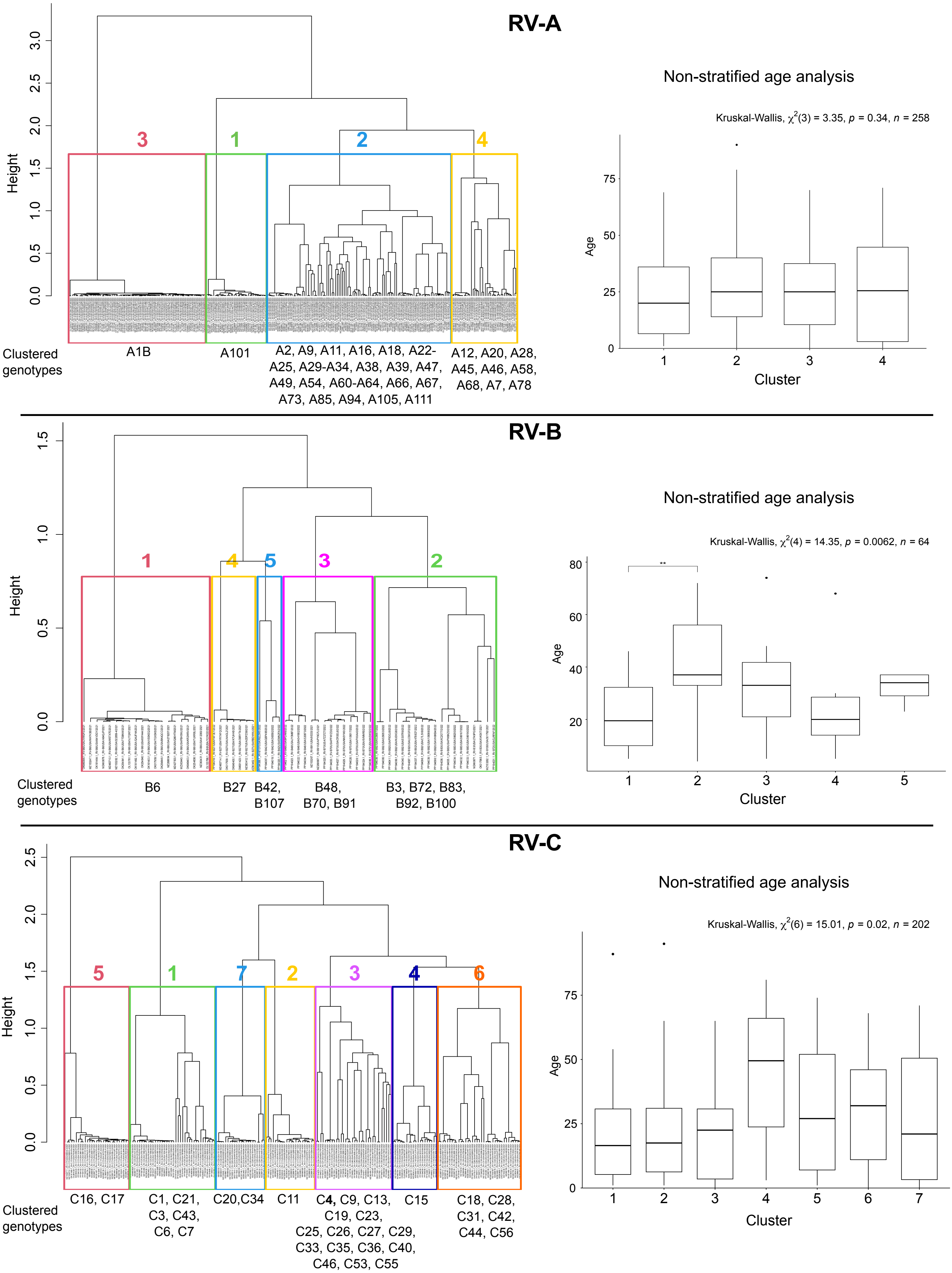
