## Supplementary Figure 4 for "Genomic epidemiology and evolution of rhinovirus in western Washington State, 2021-22"

**Supplementary Figure 4. Phylodynamic analysis of prevalent RV genotypes.**

Maximum clade credibility tree (MCCT) for prevalent RV genotypes. The inference was performed with complete genome of the sequences from Washington State dating from 2021 and 2022 together with genomes available in public databases with full collection date. The MCCT has branch lengths in time units depicted on the time scale. The time in years of the most recent common ancestors is indicated in each node. The uncertainty (95% highest posterior density intervals or 95% HPD) for the node times is indicated with blue bars. Color in branches denote the node posterior probabilities. The evolutionary rate with the 95%HDP is indicated below the name of each genotype. Clades of Washington State (USA-WA) sequences from 2021 and beyond with common ancestors dating after 2020 are collapsed into triangle. The number of collapsed sequences is indicated at the right of the triangle.

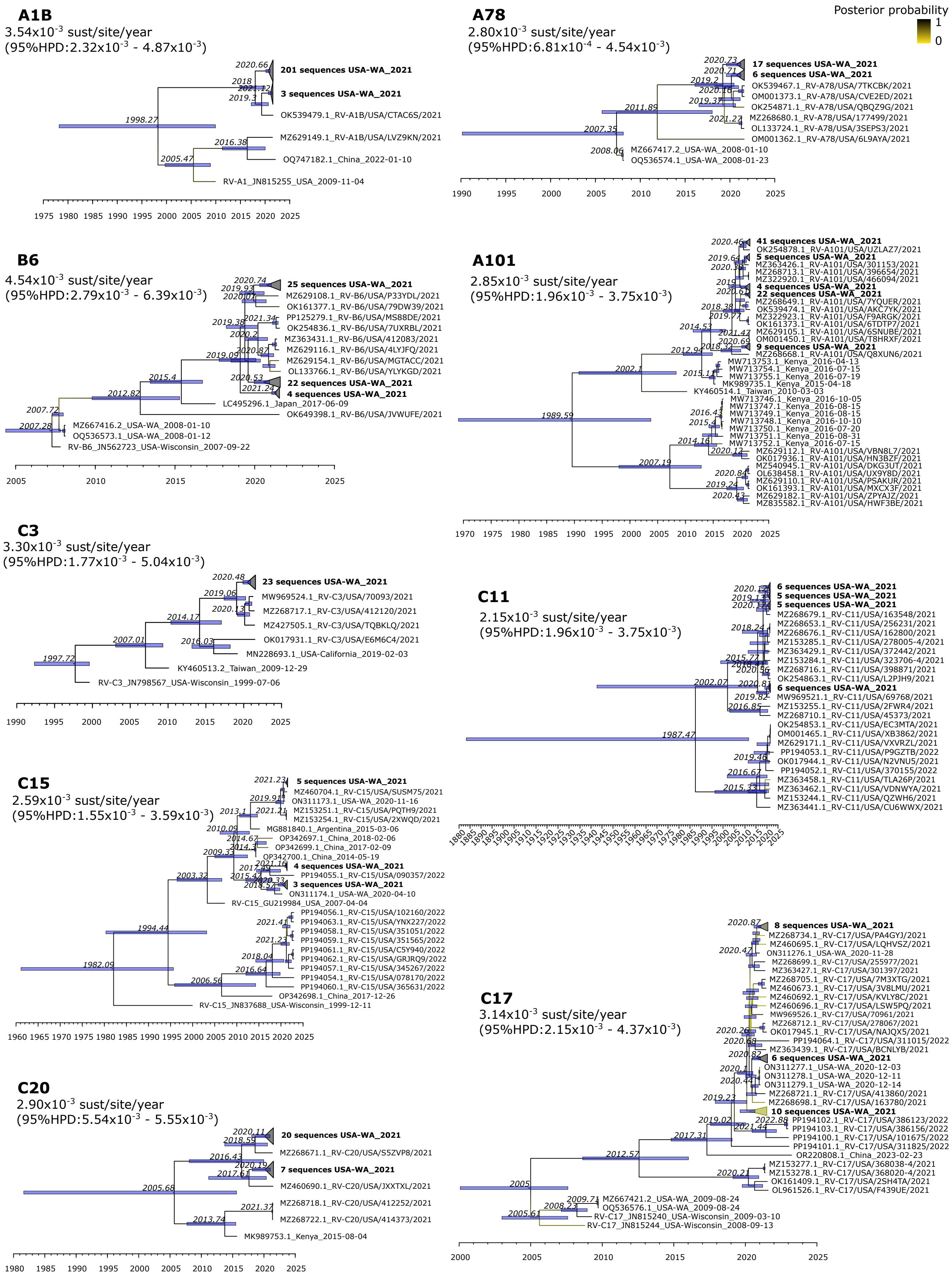
