## Supplementary Figure 5 for "Genomic epidemiology and evolution of rhinovirus in western Washington State, 2021-22"

**Supplementary Figure 5. Shannon entropy of the RV polyprotein sequence for prevalent genotypes.** Genomes from 2021 and 2022 in Washington State together with all the genomes from the same genotype available in public databases were aligned, trimmed according to the polyprotein open reading frame, and translated with the universal genetic code. Bar plots show the Shannon entropy value of each position of the entire polyprotein. Grey columns denote values below 1, meaning conserved positions. Red columns denote values above 1.

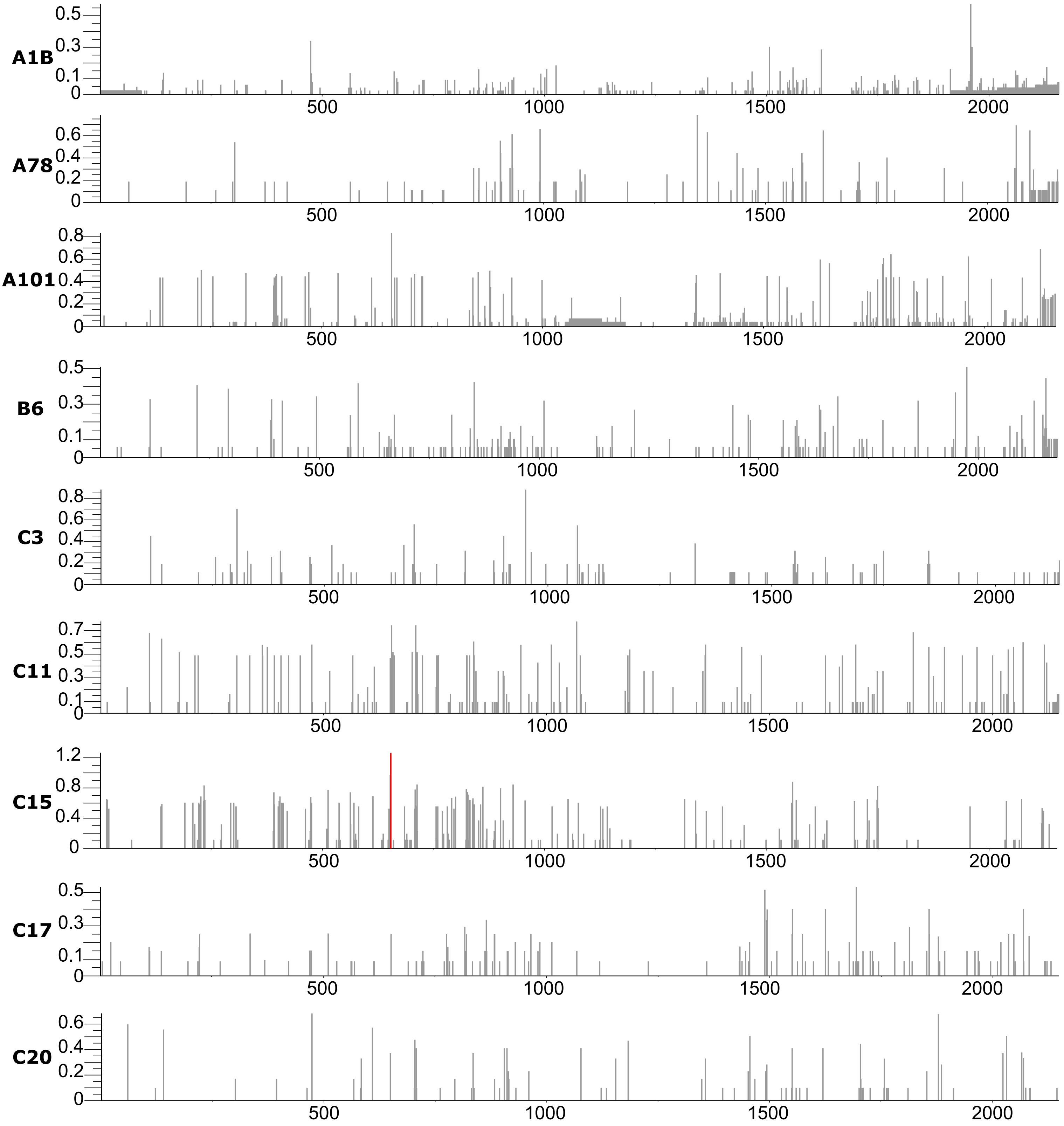
