## Supplementary Figure 6 for "Genomic epidemiology and evolution of rhinovirus in western Washington State, 2021-22"

**Supplementary Figure 6. Rhinovirus genomes available in NCBI GenBank after our study in Puget Sound region, WA.**
Bar plots show the cumulative number of genomes of each RV genotype available in NCBI after our study in Puget Sound region (database reviewed in September 2023). In yellow the number of genomes per genotype before our study is indicated and in green the number of genomes per genotype we have sequenced and publicly shared is highlighted.

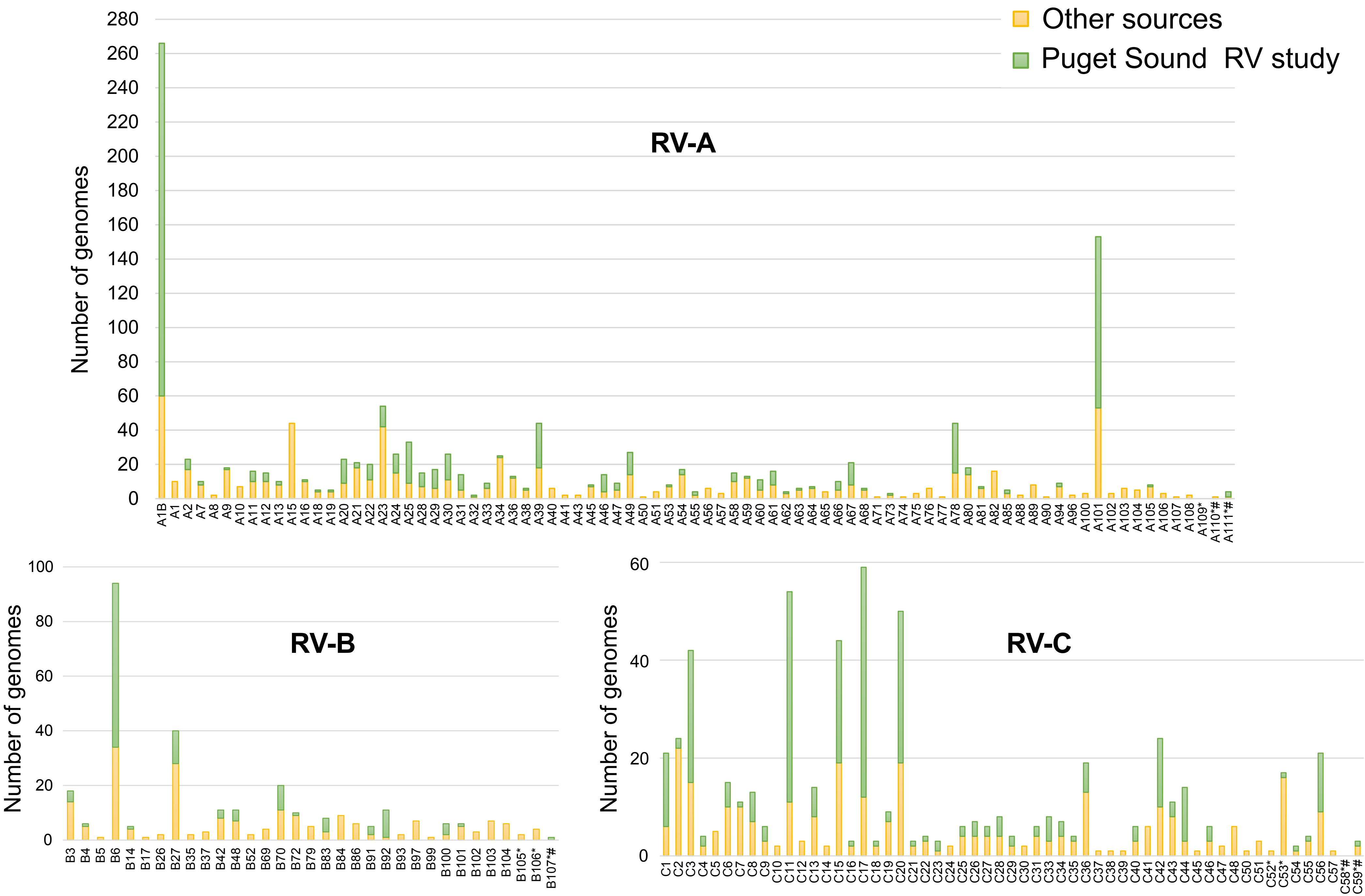


* genotype not confirmed in ICTV (https://ictv.global/report/chapter/picornaviridae/picornaviridae/enterovirus)

### genotype not confirmed by the Picornaviridae Study Group (https://www.picornastudygroup.com/)
